# The biological clock of multimorbidity: temporal dynamics of disease co-occurrence in primary care

**DOI:** 10.64898/2026.06.15.26355655

**Authors:** Jon Sánchez-Valle, Carme Zambrana, Alejandro Navarro-Martínez, Felipe Xavier Costa, Luis Rocha, Davide Cirillo, Concepción Violán, Alfonso Valencia

## Abstract

Multimorbidity is the dominant clinical reality of primary care, yet the temporal dynamics governing when and how persistent comorbidity associations emerge remain poorly characterised. Most large-scale comorbidity studies adopt a single observation window after an index diagnosis, implicitly assuming that associations detectable at one year are equally detectable at five. Using 11 years of electronic health records from 5,821,197 individuals in Catalan primary care, we applied a matched cohort design across nine complementary follow-up windows – five cumulative (0-1 to 0-5 years) and four conditional (1-2 to 4-5 years) – to 1,315 index diseases, identifying 144,030 significant directed comorbidity associations in the five-year network. We found that 60.1% of these associations required at least three years of follow-up and were undetectable in shorter-window analyses, demonstrating that observation window length is a primary determinant of which comorbidities can be observed. To organise this temporal heterogeneity, we introduce the biological clock of multimorbidity: a two-dimensional framework that positions ICD-10 disease categories according to their rates of cumulative signal attenuation and the persistence of conditional risk. This framework identifies four reproducible temporal patterns (episodic, chronic stable, chronic progressive, and transient-persistent) that are robust under bootstrap resampling, leave-one-disease-out sensitivity analysis, and alternative clustering approaches. The biological clock is systematically modulated by sex, with Blood/Immune and Musculoskeletal disorders showing the largest sex differences in temporal dynamics. Network analysis identified 19 disease “initiators” that generate broad downstream comorbidity burdens and 21 “sinks” representing convergent endpoints of multiple disease trajectories. Comparison with hospital-based Danish data from 6,909,676 individuals showed that shared associations were 2.7-fold enriched over chance expectation (hypergeometric test, p<10^-300^) and showed moderate concordance of effect sizes (Spearman ρ=0.460), confirming that the comorbidity structure identified here reflects genuine, generalisable signal; nonetheless, only 3.6% of primary care associations were replicated in the hospital network, indicating that the two settings capture largely complementary segments of the disease co-occurrence landscape. Together, these findings establish the observation window length as a principal design parameter in EHR-based multimorbidity research and the biological clock as a framework for understanding how and over what timescale disease associations emerge, persist, and resolve.

## Introduction

Multimorbidity – the co-occurrence of two or more chronic conditions in the same individual – has become the predominant clinical reality of primary care. In high-income countries, more than half of adults aged 65 and older live with multiple concurrent conditions, and the burden is rising in younger age groups ^1–4^. Patients with multimorbidity accumulate more medications, experience more hospitalisations, and show worse functional outcomes than those with single conditions ^5–7^, while simultaneously falling outside the eligibility criteria of most clinical trials and guidelines ^8–10^. Primary care is both where multimorbidity is most prevalent and where it is first detected and managed, making it the natural context for population-level characterisation of disease co-occurrence^11–13^. Large-scale electronic health record (EHR) systems now provide the data infrastructure to study multimorbidity at previously unattainable resolution and scale ^14^.

Network-based approaches to disease co-occurrence have emerged as a powerful framework for characterising multimorbidity at the population scale. The foundational work of Jensen *et al*. and Westergaard *et al*. using Danish national hospital registry data ^15,16^ established that disease co-occurrence is non-random and exhibits reproducible population-level structure. Recent analyses have extended this framework to multilayer representations and to primary care settings, examining age-, sex-, and ethnicity-stratified subgroups ^17,18^, including studies using the SIDIAP database in Catalonia ^19–24^. One fundamental but underexplored limitation of comorbidity studies is their dependence on the choice of follow-up window – the observation period after index diagnosis during which subsequent diseases are ascertained ^25^. Similarly, the largest differences between comorbidity networks arise from the source of the information ^26^. Existing studies employ observation windows ranging from contemporaneous co-occurrences to the entire available follow-up period, a heterogeneity that renders cross-study comparisons ambiguous ^15–17,27,28^, yet its impact on comorbidity detectability has never been systematically quantified. It remains unknown whether window choice changes which comorbidities are detected, how strongly, or whether these effects differ across disease categories. Equally uncharacterised is the conditional temporal structure of comorbidity risk: the probability of developing a secondary condition among patients who have not yet developed it in earlier intervals – a dimension invisible to conventional cumulative analyses and unexplored as an independent axis of temporal variation.

The choice of observation window also has direct consequences for sex-stratified analyses: sex differences in multimorbidity are well documented in disease prevalence and incidence ^29–32^, and Westergaard *et al*. showed that the temporal ordering of some disease pairs is reversed between women and men ^16^. Whether sex differences extend to the temporal dynamics of comorbidity – whether the rate at which comorbidity signal attenuates and the persistence of late-onset conditional risk differ between women and men – remains unexplored.

Here, we address these gaps using 11 years of primary care EHR data from 5,821,197 individuals in Catalonia, Spain (SIDIAP, 2008-2018), applying five cumulative and four conditional follow-up windows to characterise the complete temporal signature of each comorbidity association. We show that 60.1% of significant disease associations require at least three years of follow-up to become detectable, demonstrating that observation window length is a primary determinant of multimorbidity network structure.

To organise this temporal heterogeneity, we introduce the biological clock of multimorbidity: a two-dimensional space that positions ICD-10 disease categories by their rate of signal attenuation and the persistence of conditional risk, defining four temporally coherent patterns. We further show that the biological clock is systematically modulated by sex, that network analysis identifies a small set of disease initiators and sinks linking temporal dynamics to network position, and that comparison with hospital-based Danish data validates the generalisability of the network while revealing that primary care and hospital comorbidity networks capture largely complementary segments of the disease co-occurrence landscape.

## Results

### Observation window length determines comorbidity detectability

Across all nine follow-up windows, 146,591 significant directed comorbidity associations were identified in at least one window. The largest group comprised 72,495 long-window-only pairs (49.5%) (associations detectable only in cumulative windows spanning three or more years, with no signal in the 0-1-year window and no conditional risk in the non-overlapping intervals), representing comorbidity relationships that are systematically invisible to studies using short follow-up windows. The second-largest group comprised 45,495 omnipresent pairs (31.0%), associations detectable in all nine windows simultaneously, with residual conditional risk persisting throughout the observation period. The remaining pairs fell into smaller but informative classes: 15,645 (10.7%) showed late cumulative signal combined with persistent conditional risk; 7,908 (5.4%) were persistent across all cumulative windows but with only partial conditional signal; 2,445 (1.7%) were persistent cumulatively without any conditional risk; and only 157 (0.1%) were early transient (detectable only in the 0-1-year window) confirming that extending follow-up primarily reveals previously undetectable associations. In summary, 60.1% of associations (88,140 pairs) were late-emerging, detectable only with three or more years of follow-up, while 38.1% (55,848) were robust to window choice, and only 0.8% (1,100) were transient or conditional-only. This polarisation was reflected in the cumulative networks: from 56,084 pairs at 0-1 years to 144,030 at 0-5 years, a 2.6-fold increase (Fig. 1).

**Figure 5.**
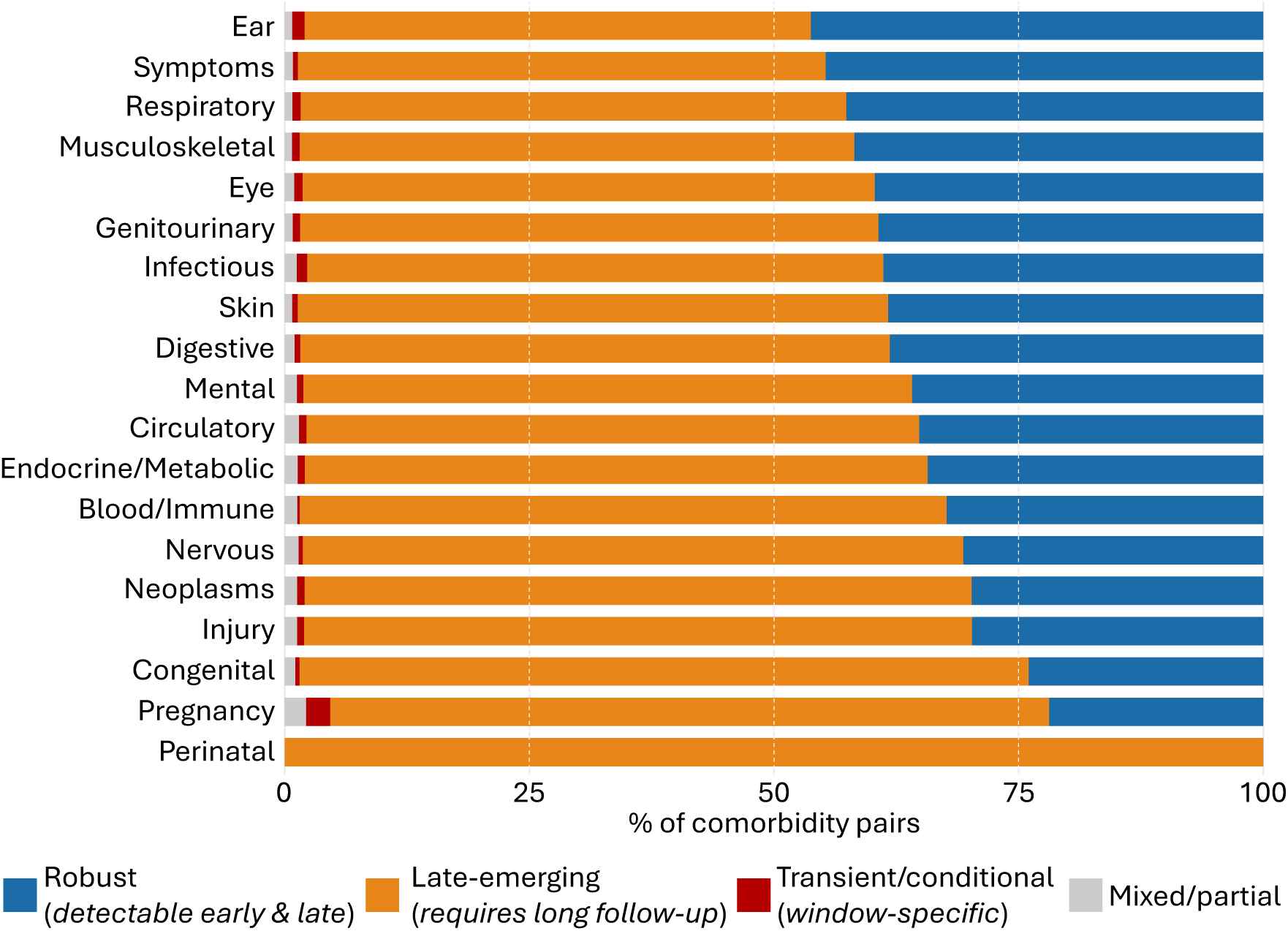
Temporal robustness of comorbidity associations by ICD-10 disease category. Stacked bar chart showing, for each ICD-10 category of the index disease, the proportion of comorbidity pairs belonging to three temporal detection groups: blue = robust (detectable regardless of follow-up window choice, encompassing omnipresent, persistent cumulative, and persistent cumulative with partial conditional pairs); orange = late-emerging (requiring cumulative follow-up of three or more years, with or without additional conditional signal); red = transient or conditional only (detectable exclusively in specific time windows, including early transient, conditional only, and early cumulative with late conditional pairs). Grey = mixed or partial patterns. Categories are ordered by decreasing proportion of robust associations. Total pairs analysed: 146,591 directed disease pairs with significant co-occurrences in at least one of the nine follow-up windows. Analysis performed for the combined population (both sexes).

The distribution of temporal classes was highly structured across ICD-10 disease categories (Fig. 1; Supplementary Figure 1). Ear disorders showed the highest proportion of omnipresent associations (40.7%), followed by Musculoskeletal (36.3%), Symptoms (36.2%), Respiratory (35.3%), and Eye (33.8%) disorders – predominantly chronic, recurrent, or organ-specific conditions that generate comorbidity signals detectable regardless of the chosen follow-up window. In contrast, Perinatal conditions showed the most extreme long-window dependence: all eight significant Perinatal pairs were invisible in the 0-1-year window and required extended follow-up to emerge, consistent with neurodevelopmental, metabolic, and cardiovascular sequelae that typically manifest years after the initial perinatal episode. Congenital disorders showed a similar pattern (65.2% long-window-only, 17.4% omnipresent). Nervous system (58.6% long-window-only, 24.9% omnipresent), Blood/Immune (57.8%, 24.9%), and Neoplasms (55.8%, 24.6%) showed high long-window dependence despite substantial omnipresent fractions, the latter reflecting the long latency of oncological sequelae and treatment complications. Category-pair analyses revealed additional structured patterns: Ear disorders as secondary disease showed consistently high omnipresent proportions (40-60%) across all index categories, while Perinatal and Congenital conditions as index diseases showed 100% long-window dependence across all secondary categories (Supplementary Figures 2-5).

An extended temporal taxonomy combining cumulative and conditional patterns across all 419,995 pairs with data in all nine windows revealed additional heterogeneity not captured by the three-group summary, including 8,604 pairs with an amplifying cumulative signal (RR increasing with extended follow-up) and 8,978 pairs with a late-emerging cumulative signal combined with persistent conditional risk, patterns invisible to analyses restricted to cumulative windows alone (Supplementary Figure 6). Ultrametric closure analysis confirmed that the choice of temporal window affects not only the number of detected associations but also the qualitative structure of multi-step disease trajectories, with neoplasm and congenital conditions emerging as trajectory origins exclusively in the 5-year window (Supplementary Figure 7; Supplementary Text).

### Temporal heterogeneity of disease associations

Among the 144,030 significant directed associations in the 0-5-year network, 55,848 were consistently detected across all five cumulative windows. For these persistent pairs, the posterior relative risk showed systematic attenuation over time (median change −7.9%, IQR −15.1% to −1.0%), with 85.5% showing less than 20% change and only 0.5% strengthening. The rate of attenuation differed markedly across ICD-10 categories (Kruskal-Wallis χ²=1,360.7, df=17, p<2.2×10^-16^). Pregnancy (−0.060), Symptoms (−0.053), and Skin (−0.049) showed the fastest attenuation, consistent with acute and episodic disease relationships, whereas Congenital (−0.027), Neoplasms (−0.027), and Endocrine/Metabolic (−0.025) disorders showed the slowest, consistent with chronic progressive diseases maintaining stable comorbidity associations across the observation period.

Complementing the cumulative attenuation analysis, we assessed whether comorbidity risk persists in patients who have not yet developed the secondary condition. Among 436,863 disease pairs with data in all four conditional windows, 59,644 (13.7%) showed persistent conditional risk, significant associations in both early (years 1-2 or 2-3) and late (years 3-4 or 4-5) windows, and 5,646 (1.3%) showed late-emerging conditional risk exclusively in later windows. The proportion of comorbidities with persistent conditional risk varied substantially across categories: Ear (21.4%), Musculoskeletal (18.2%), and Respiratory (17.1%) disorders showed the highest persistent conditional risk, whereas Perinatal (0%), Congenital (3.5%), and Pregnancy (5.0%) showed the lowest.

These two dimensions, rate of cumulative attenuation and proportion of pairs with persistent conditional risk, were largely independent across categories, defining a two-dimensional space of temporal comorbidity behaviour that motivates the biological clock framework introduced in the next section (Fig. 2A).

**Figure 2.**
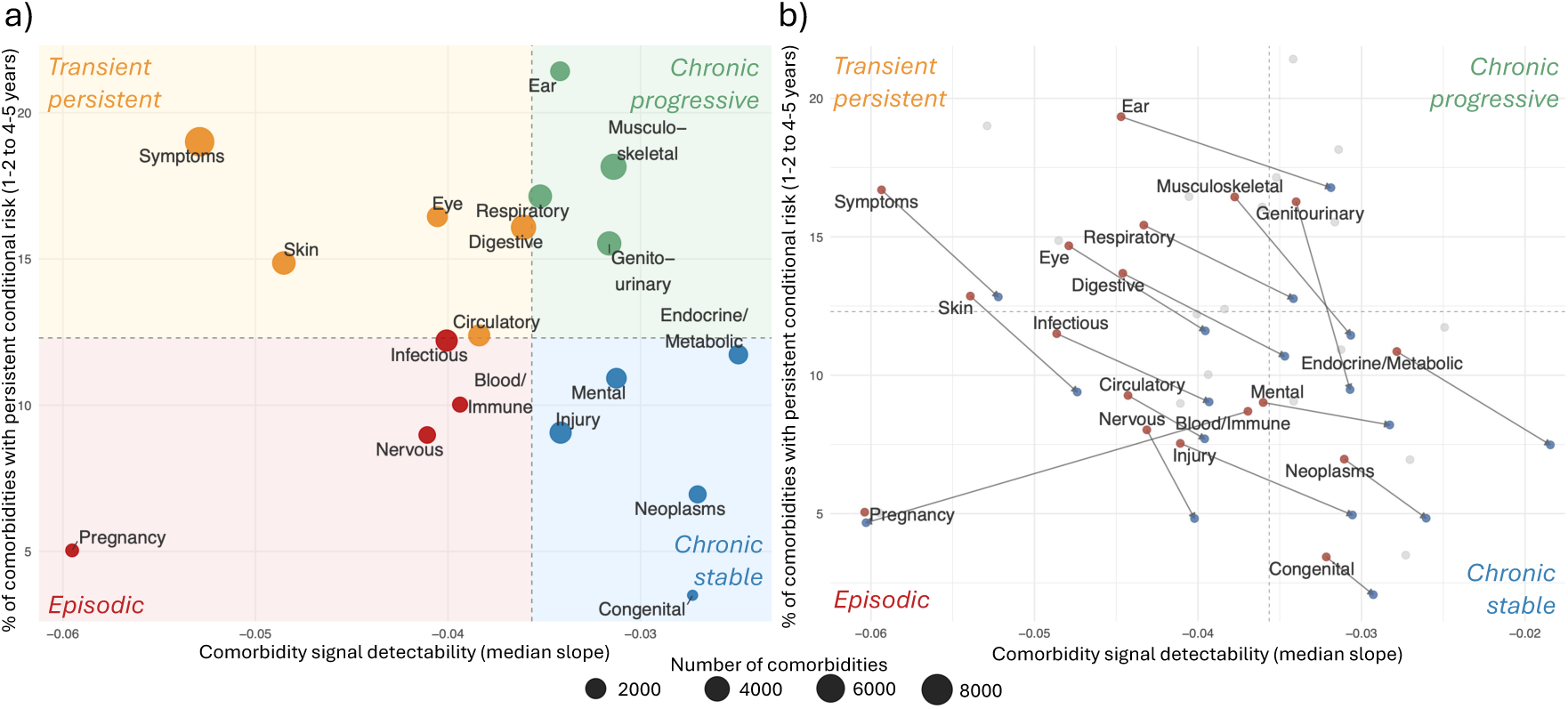
The two-dimensional biological clock of multimorbidity and its sex-specific variation. **a)** Two-dimensional temporal dynamics space. Each point represents an ICD-10 disease category. The x-axis shows the comorbidity signal detectability, calculated as the median-weighted slope of the posterior RR across the five cumulative follow-up windows (0-1 to 0-5 years) for persistent comorbidity pairs. The y-axis shows the proportion of comorbidity pairs within each category that display persistent conditional risk, defined as significant associations in both the early (years 1-2 or 2-3) and late (years 3-4 or 4-5) conditional windows. Quadrant boundaries are defined by the median values of both dimensions. Colours indicate biological clock patterns: red = episodic, green = chronic progressive, blue = chronic stable, and orange = transient-persistent. Point size is proportional to the number of persistent comorbidity pairs. **b)** Sex-specific variation in the two-dimensional biological clock. Grey points indicate the position of each ICD-10 category in the combined population, while coloured points indicate sex-specific estimates (red = women; blue = men). Arrows connect the female and male positions for each category, indicating the direction and magnitude of sex-related displacement within the temporal dynamics space. The x-axis represents the rate of RR attenuation, and the y-axis the proportion of comorbidity pairs with persistent conditional risk. Categories that show larger displacements exhibit greater sex differences in the temporal behaviour of multimorbidity associations.

### The biological clock of multimorbidity

A central question raised by the temporal heterogeneity described above is whether disease associations differ systematically in how their signals emerge, attenuate, and persist over time (and whether this temporal behaviour is itself structured across disease categories). Combining the rate of cumulative signal attenuation and the proportion of pairs with persistent conditional risk positions ICD-10 disease categories in a two-dimensional temporal space within the 1-5-year observation range, the biological clock of multimorbidity (Fig. 2A). Four reproducible temporal patterns emerge. Episodic categories (Pregnancy, Blood/Immune, Nervous, Infectious) show fast cumulative decay and low persistent conditional risk, consistent with disease relationships concentrated in the acute post-diagnosis period. Chronic stable categories (Mental, Injury, Endocrine/Metabolic, Congenital, Neoplasms) exhibit slow decay and low conditional persistence, reflecting durable associations largely independent of observation window length within the range studied. Chronic progressive categories (Musculoskeletal, Genitourinary, Ear, Respiratory) combine slow decay with high persistent conditional risk, indicating that significant comorbidity risk remains in patients who have not yet developed the secondary condition, even years after index diagnosis. Transient-persistent categories (Circulatory, Digestive, Eye, Skin, Symptoms) show fast cumulative decay combined with high conditional risk; rather than forming a fully discrete class, these categories are best understood as occupying a transitional zone between episodic and chronic progressive patterns, where cumulative signal fades quickly but underlying disease susceptibility persists.

These patterns are defined at the category level, reflecting the median behaviour of hundreds of comorbidity pairs within each ICD-10 chapter. Two examples illustrate the key distinctions. Alcohol-related disorders → alcoholic liver cirrhosis (F10→K70, chronic stable) shows a cumulative RR stable between 25.5 and 28.3 across all five years with significant conditional risk in all four annual intervals (RR 27.5 at year 2 to 18.3 at year 5), consistent with a persistent underlying disease process operating regardless of observation window. Acute sinusitis → chronic sinusitis (J01→J32, chronic progressive) shows declining cumulative RR from 29.8 at year 1 to 12.5 at year 5 with persistent conditional risk maintained throughout (RR 13.0 at year 2, 6.4 at year 5), consistent with progressive inflammatory remodelling in which susceptibility to recurrence persists even as the cumulative signal weakens. The episodic and transient-persistent patterns are illustrated by A63→A64 and R06→J96, respectively (Fig. 3; Supplementary Table 1).

**Figure 3.**
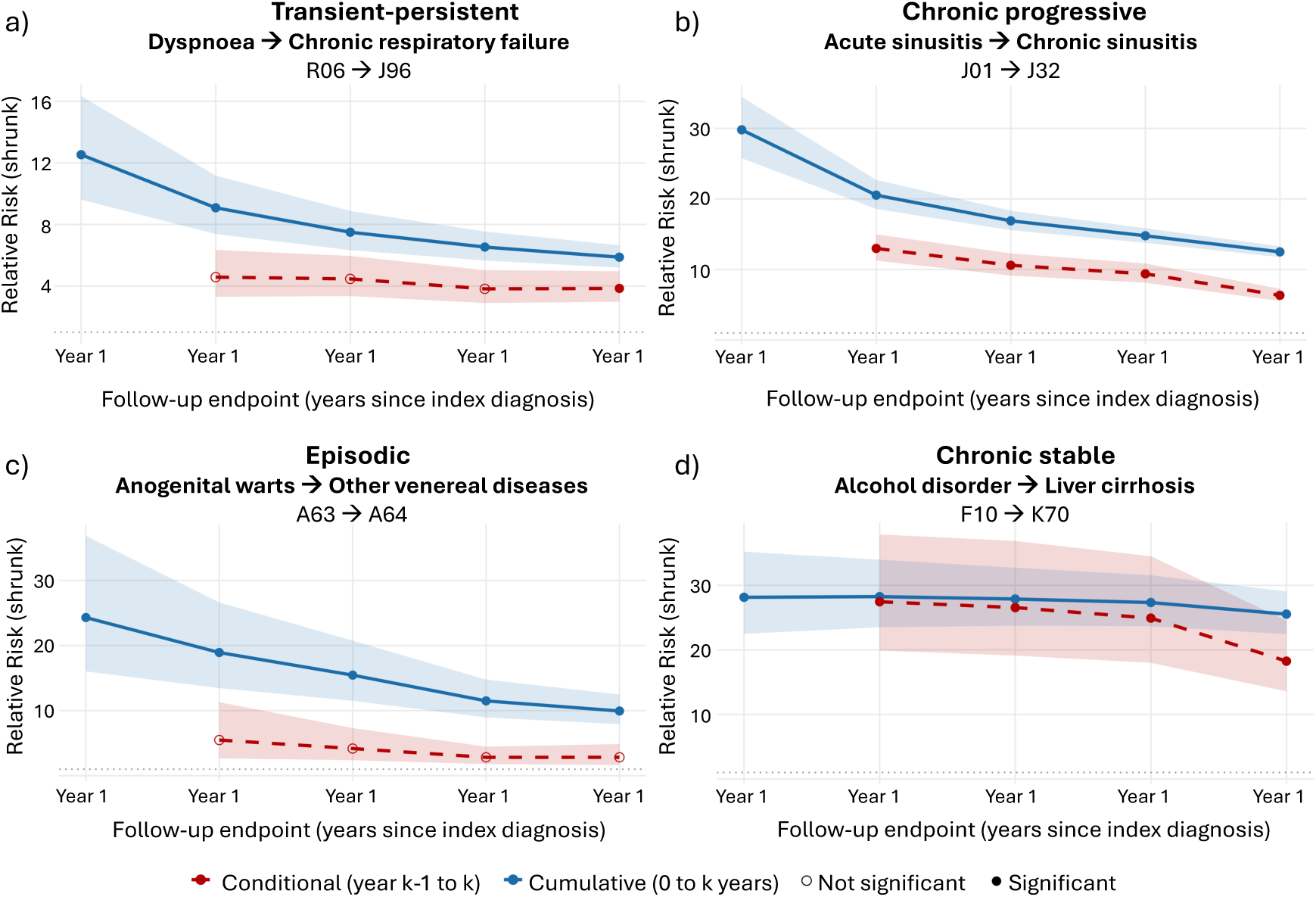
Paradigmatic examples of the four biological clock patterns. Each panel shows the temporal trajectory of one representative comorbidity pair across five annual endpoints. Blue solid lines show cumulative RR estimates (0 to endpoint year); red dashed lines show conditional RR estimates (risk in the year preceding the endpoint, given no prior occurrence of the secondary disease). Both lines share the same x-axis endpoint, allowing direct comparison of cumulative and conditional risk at each time point. Filled circles indicate statistically significant associations (LFSR < 0.05, lower bound of the 95% credible interval ≥ 1.01, and ≥ 100 exposed patients); open circles indicate associations that do not meet these criteria. Shaded ribbons indicate 95% credible intervals and are shown for all timepoints regardless of significance. For the transient-persistent example (R06→J96, dyspnoea → chronic respiratory failure), conditional windows in years 2-4 do not meet the ≥100 exposed-patients threshold, despite a significant LFSR and a credible interval lower bound > 1.01; the year 5 conditional window meets all significance criteria. The transient-persistent pattern is primarily a category-level property, reflecting a combination of rapid cumulative decay and a high proportion of transient-persistent conditional risk across the category; individual pairs within transient-persistent categories may exhibit transient-persistent rather than strictly transient-persistent conditional trajectories.

Clock positions were robust under pair-level bootstrap resampling across most categories (Supplementary Figure 8; Supplementary Table 2). Chronic progressive categories showed the highest stability: Musculoskeletal and Genitourinary retained their quadrant assignment in 100% of bootstrap replicates, followed by Ear (90.2%) and Respiratory (71.0%). Episodic categories also showed high stability: Pregnancy (100%) and Blood/Immune (95.7%), while Nervous (53.0%) and Infectious (41.3%) showed moderate retention, consistent with their proximity to the cumulative attenuation boundary. Chronic stable categories were similarly robust: Mental (99.6%), Injury (96.1%), Congenital (92.6%), and Endocrine/Metabolic (87.6%) all retained their classification in the large majority of replicates. Leave-one-index-disease-out analysis confirmed these results for the most stable categories: Genitourinary and Musculoskeletal retained their chronic progressive classification regardless of which index disease was excluded (100% retention), and Ear and Respiratory showed retention of 92.3% and 78.4%, respectively. Transient-persistent categories (Circulatory, Digestive, Eye, Skin, Symptoms) showed lower bootstrap retention (0%), reflecting their position near the cumulative attenuation boundary (their bootstrap confidence intervals for median slope (e.g. Digestive: 95% CI −0.037 to −0.034) spanned the median attenuation boundary (−0.036), causing them to alternate between quadrants across replicates). K-means clustering (k=4) agreed with the median-split classification for 12 of 18 categories (66.7%), with concordance highest for Chronic progressive (4/4) and Chronic stable (5/5), and lowest for Episodic (1/4) and Transient-persistent (2/5) (where k-means assigned Skin and Symptoms to a transient-persistent cluster and Circulatory, Digestive, and Eye to Chronic progressive) consistent with the gradient interpretation.

Analysis at the ICD-10 subcategory level revealed substantial within-category heterogeneity, indicating that chapter-level clock positions represent central tendencies masking clinically relevant variation at finer diagnostic resolution (Supplementary Figure 9; Supplementary Table 3). Endocrine/Metabolic disorders showed the broadest internal diversity (slope range = 0.10 RR units/year; persistent conditional risk range = 25 percentage points), spanning all four clock quadrants: glucose dysregulation disorders (E15-E16) occupied the episodic quadrant (slope = −0.118, persistent risk = 3.9%), while obesity and hyperalimentation disorders (E65-E68) showed the highest persistent conditional risk in this chapter (slope = −0.016, persistent risk = 28.8%, chronic progressive). Mental disorders showed comparable heterogeneity (slope range = 0.056; persistent risk range = 18.6 percentage points), also spanning all four quadrants: organic mental disorders (F00-F09) were episodic (slope = −0.077, persistent risk = 3.0%), whereas neurotic and stress-related (F40-F48) and mood disorders (F30-F39) occupied the chronic progressive quadrant (persistent risk = 18.4% and 15.4% respectively). Within Circulatory disorders, ischaemic heart diseases (I20-I25) showed a transient-persistent profile (persistent risk = 15.5%), in contrast to cerebrovascular diseases (I60-I69), which were episodic (slope = −0.065, persistent risk = 4.9%). Ear disorders showed the narrowest internal variation (slope range = 0.025; persistent risk range = 10.4 percentage points), with all four subcategories occupying the chronic progressive or transient-persistent quadrants, confirming the robustness of the chapter-level classification. Pregnancy subcategories showed the most extreme slope heterogeneity (range = 0.33 RR units/year), driven by the strongly episodic signal of maternal care disorders (O30-O48; slope = −0.352), consistent with the acute and time-limited nature of obstetric complications.

### Sex-specific temporal dynamics

The biological clock was broadly conserved between women and men but showed systematic differences in both temporal dimensions (Fig. 2B). Women showed faster cumulative attenuation than men across most categories, including Circulatory (−0.044 vs −0.040), Respiratory (−0.043 vs −0.034), and Skin (−0.054 vs −0.047). The principal exception was Blood/Immune disorders, where men showed markedly faster attenuation (−0.060 vs −0.037), consistent with the predominance of chronic autoimmune conditions (notably lupus, rheumatoid arthritis, and Sjögren syndrome) in women versus acute haematological and reactive immune conditions in men. Women showed higher persistent conditional risk across nearly all categories, with the largest differences in Genitourinary (16.3% vs 9.5%) and Musculoskeletal (16.4% vs 11.4%).

These temporal differences deepen a pattern of sex-specific disease burden already visible in static measures. Sex-biased prevalence was a widespread feature of the disease landscape: 429 of 1,498 ICD-10 diseases were significantly more prevalent in women and 332 in men, with women-biased diseases enriched in genitourinary, musculoskeletal, and endocrine/metabolic categories, including obesity (E66), depressive episode (F32), and asthma (J45), and men-biased diseases enriched in neoplasms and circulatory disorders, including type 2 diabetes (E11) and chronic ischaemic heart disease (I25) (Fig. 4A-B). Sex differences were equally widespread in age at first recorded diagnosis: type 2 diabetes was diagnosed 4.78 years later in women despite being more prevalent in men, and schizophrenia showed one of the largest diagnostic delays in women (10.21 years), while alcohol- and tobacco-related disorders were both more prevalent and diagnosed later in men (3.41 and 5.5 years, respectively). At the category level, significantly later diagnosis in women was observed for injury/poisoning, circulatory, endocrine/metabolic, musculoskeletal, and mental disorders, while blood/immune and genitourinary diseases were diagnosed significantly later in men (Supplementary Table 4). These patterns were largely concordant with Danish hospital data, supporting shared biological components of sex-differential disease timing (Supplementary Text). The temporal framework thus reveals a layer of sex-specific comorbidity structure that prevalence-based analyses alone do not capture.

**Figure 4.**
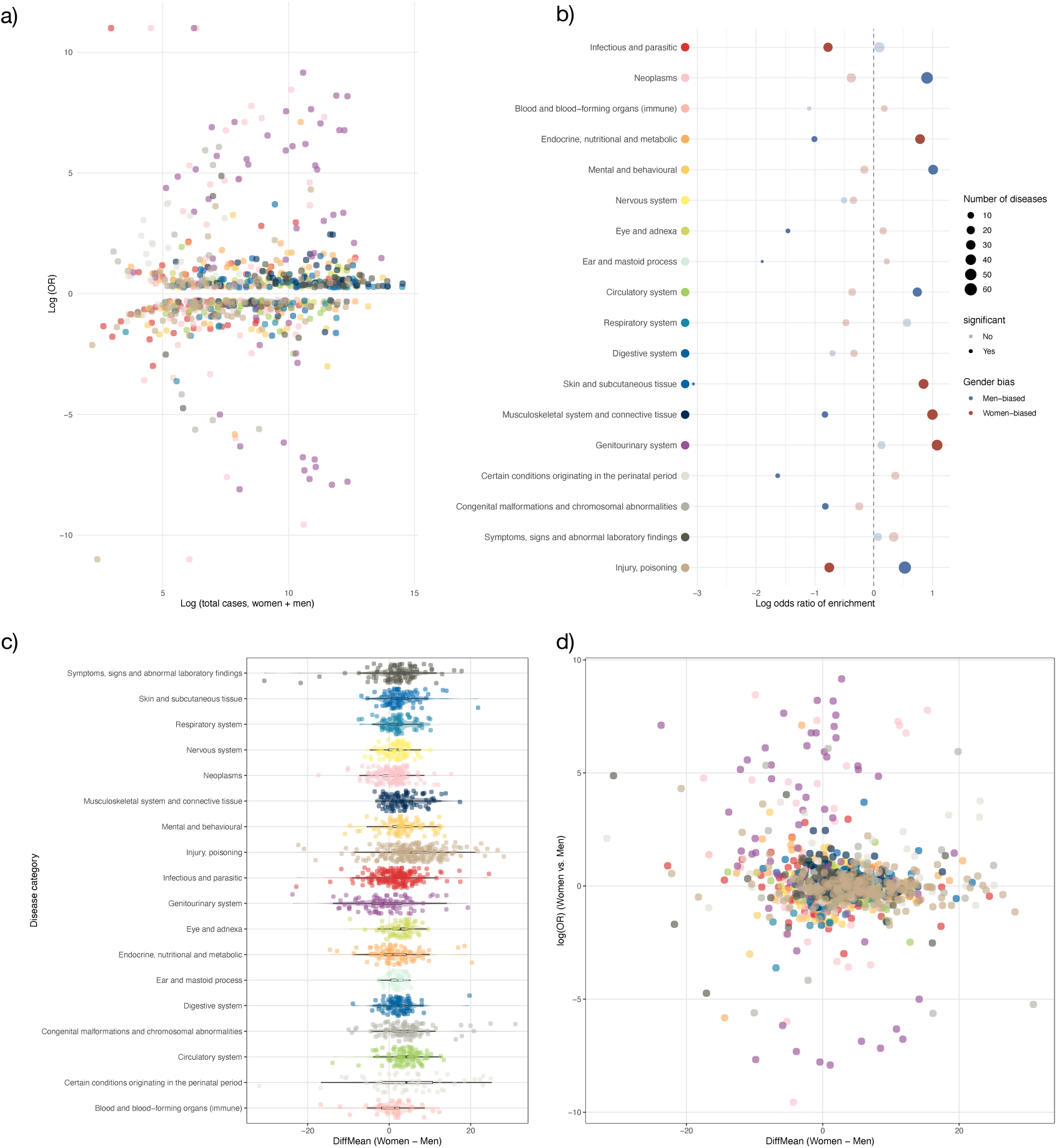
Sex differences in disease prevalence and age at first diagnosis across 1,498 ICD-10 diseases in Catalonia (2008–2018). (a) Log-odds ratio (OR) of disease prevalence in women versus men, plotted against the log of the total number of diagnosed individuals. Each point represents one disease; colours indicate ICD-10 category. Only diseases meeting significance thresholds are shown (FDR ≤ 0.05 and OR ≥ 1.3 or ≤ 1/1.3). (b) Enrichment of ICD-10 categories among diseases with significantly higher prevalence in women (red) or men (blue). The x-axis shows the log OR of enrichment; dot size reflects the number of sex-biased diseases per category; filled dots indicate significant enrichment after FDR correction. (c) Disease-specific mean age differences at first recorded diagnosis (women − men, years) across ICD-10 categories. Each point represents one three-digit ICD-10 code. Positive values indicate later diagnosis in women; negative values indicate later diagnosis in men. (d) Joint distribution of sex differences in diagnostic age and disease prevalence (log OR, women versus men), illustrating the heterogeneity and partial independence of the two dimensions across the disease landscape.

The comorbidity networks showed asymmetric overlap: women had a larger network (110,690 associations vs 79,772 in men), with 85.2% of men’s associations present in the women’s network but only 61.4% of women’s associations present in the men’s network, reflecting the broader ambulatory disease spectrum in women. Shared associations were enriched in chronic progressive transitions, particularly Ear→Ear (OR=17.3) and Respiratory→Ear (OR=5.4), while Genitourinary transitions were systematically under-represented (Fig. 5A). Comparing the two networks directly, transitions involving endocrine, genitourinary, and injury-related diseases were significantly more prevalent in women, while Mental→Mental (OR=0.57), Circulatory→Circulatory (OR=0.69), and Mental→Circulatory (OR=0.69) transitions were more prevalent in men, reflecting well-documented sex differences in cardiovascular and substance-related mental health comorbidity (Fig. 5B).

**Figure 5.**
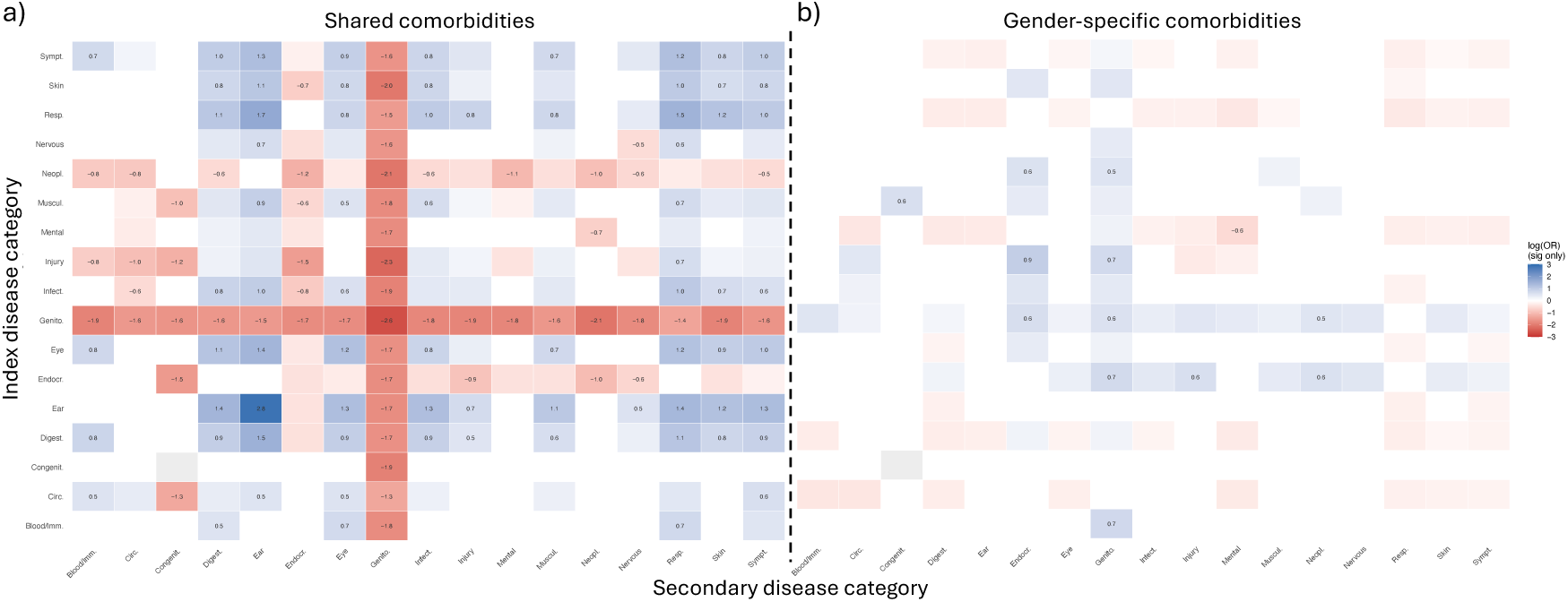
Sex differences in comorbidity network structure (window 0-5 years). **a)** Enrichment of ICD-10 category pairs among shared comorbidities (present in both women and men networks) versus sex-specific pairs. Blue = overrepresented among shared pairs; red = underrepresented. **b)** Differential representation of ICD-10 category pairs in women versus men. Blue = overrepresented in women; red = overrepresented in men. Both panels: Fisher’s exact test with Benjamini-Hochberg correction; only significant cells shown (FDR ≤ 0.05); grey = not significant or no data. Pregnancy and Perinatal conditions are excluded as index diseases because they are absent or negligible in the men’s network.

Sex-based directional reversals were rare (<2.5% in any category) but concentrated in Blood/Immune (2.4%), Skin (2.0%), and Symptoms (1.2%). The most clinically coherent examples were gallstones → bile duct obstruction (K80→K42; A→B preferred in women, RR=3.9, θ=0.59; reversed in men, RR=1.5, θ=0.56), consistent with sex differences in biliary disease presentation; and type 2 diabetes → gout (E11→M10; A→B preferred in women, RR=2.5, θ=0.62; reversed in men, RR=1.4, θ=0.52), consistent with the earlier onset of gout in men and different metabolic trajectories linking diabetes and gout across sexes. Directional stability across temporal windows was high in both sexes, with approximately 85% of pairs maintaining their preferred direction from 0-1 to 0-5 years; reversals between windows were most common in Symptoms (13.7%), Endocrine/Metabolic (9.5%), and Infectious (8.6%) categories, consistent with short-window detection in these categories reflecting transient shared clinical contexts rather than stable directed relationships. (Supplementary Text).

### Initiator and sink roles in the comorbidity network

To identify conditions whose downstream comorbidity burden differs from their overall centrality in the network, we characterised each disease using weighted outdegree (direct downstream associations) and PageRank (recursive importance across the network), classifying diseases as initiators or sinks based on the discrepancy between these two measures. This analysis identified 19 multimorbidity initiators and 21 sinks. Initiators (conditions with high weighted outdegree relative to network PageRank) included essential hypertension (I10; z=3.99), prostate cancer (C61; z=3.48), thalassaemia (D56; z=3.42), colon cancer (C18; z=2.71), acute myocardial infarction (I21; z=2.55), and osteoporosis (M81; z=2.79), conditions that act as high-magnitude initiators of multimorbidity trajectories whose direct consequences do not themselves become network hubs. Sinks (conditions with high PageRank relative to outdegree) included hepatic failure (K72; z=-6.61), secondary malignant neoplasms (C79, C78; z=-4.63, −4.28), hepatocellular carcinoma (C22; z=-3.36), pressure ulcers (L89; z=-3.18), lower limb ulcers (L97; z=-3.18), and polyneuropathies (G62; z=-2.38), conditions that represent the convergent endpoints of multiple independent pathophysiological trajectories (Supplementary Table 5, Supplementary Figure 10).

Initiators were enriched in the chronic stable and transient-persistent categories, while sinks were enriched in the chronic progressive categories, suggesting that network role and temporal dynamics capture complementary, non-redundant dimensions of multimorbidity organisation. The initiator-sink distinction may have implications for clinical prioritisation: initiator diseases generate broad downstream comorbidity burdens whose direct consequences do not propagate further through the network, suggesting that their prevention or management could reduce a disproportionate share of downstream multimorbidity. Sink diseases represent convergent endpoints of multiple independent disease trajectories; their appearance in a patient’s record may signal the convergence of upstream processes and prompt a systematic review of the broader comorbidity context.

### Complementarity of primary care and hospital disease networks

To assess whether the comorbidity network structure identified in Catalan primary care extends beyond this specific population and setting, we compared the Catalan 0-5-year network with hospital-based Danish data from 6,909,676 individuals, restricted to 1,204 shared diseases. Despite differences in healthcare setting, follow-up window, and population composition, effect sizes among shared pairs showed moderate but consistent concordance (Spearman ρ=0.460, p<10^-265^; women: ρ=0.429; men: ρ=0.475), indicating that a conserved backbone of comorbidity associations is preserved across systems. This backbone is highly non-random: shared pairs were 2.7-fold more than expected by chance (hypergeometric test, p<10^-300^), with the enrichment consistently higher in sex-stratified analyses (women: 3.4-fold; men: 4.2-fold). Shared pairs were strongly enriched in within-category transitions: Ear (OR=19.8), Blood/Immune (OR=15.0), Mental (OR=12.2), and Circulatory (OR=7.9), reflecting associations of sufficient magnitude to be detectable in both settings. Among cross-category transitions, Digestive → Blood/Immune (OR=4.6) and Digestive → Mental (OR=3.7) were also enriched, corresponding to well-established clinicopathological relationships detectable in both ambulatory and hospital settings (Supplementary Figure 11).

Beyond this conserved backbone, structural overlap was limited: 5,109 shared pairs from 140,461 Catalan and 19,239 Danish associations (Jaccard=0.033; Table 1), with only 3.6% of Catalan associations replicated in the Danish network, though 26.6% of Danish pairs were recovered in Catalonia. This limited overlap is best understood as reflecting the distinct clinical roles of each setting rather than population differences between Catalonia and Denmark: the Danish network was enriched for Perinatal, Congenital, and Neoplasm transitions capturing high-severity trajectories under-ascertained in primary care, while transitions involving Ear disorders as secondary conditions – among the most common in Catalonia (e.g., Digestive → Ear, Genitourinary → Ear) – were entirely absent from the Danish network, consistent with the predominantly ambulatory management of ear conditions (Supplementary Figure 12). Together, these findings establish that primary care and hospital datasets capture complementary rather than redundant comorbidity landscapes.

**Table 1.** Overlap and RR concordance between Catalonia (primary care, window 0-5 years) and Denmark (hospital-based ^16^), restricted to the common disease universe (n = 1,204 diseases).

| Population | N<br>Catalonia | N<br>Denmark | N<br>shared | Jaccard | % Cat<br>in<br>Den | % Den<br>in Cat | Spearman<br>$\rho$ (log RR) | p-value |
| --- | --- | --- | --- | --- | --- | --- | --- | --- |
| Both | 140,461 | 19,239 | 5,109 | 0.033 | 3.6% | 26.6% | 0.460 | $< 10^{-265}$ |
| Women | 108,174 | 13,323 | 3,408 | 0.029 | 3.2% | 25.6% | 0.429 | $< 10^{-152}$ |
| Men | 78,689 | 12,646 | 2,869 | 0.032 | 3.7% | 22.7% | 0.475 | $< 10^{-161}$ |

Among 4,897 shared directed pairs, 57.6% showed concordant preferred directions between Catalonia and Denmark (significantly above the 50% expected by chance), indicating that the underlying temporal ordering is preserved across healthcare contexts for most shared associations. Directional reversals (n=910) were concentrated in Symptoms (n=184), Mental disorders (n=121), and Musculoskeletal (n=114), consistent with different severity ascertainment profiles. Individual reversals illustrated clinically coherent mechanisms: fatigue/malaise → lung cancer (R53 → C34) showed R53 → C34 in Catalonia (θ=0.59) but C34 → R53 in Denmark (θ=0.86), reflecting that in primary care, non-specific fatigue precedes cancer diagnosis as a prodromal symptom, whereas in hospital, the established cancer precedes fatigue as a treatment complication. These reversals do not reflect statistical noise but genuine differences in the clinical role of each setting (diagnostic in primary care, therapeutic in hospital), which determine the observable temporal ordering of co-occurring conditions. Ultrametric closure analysis confirmed this complementarity extends to multi-step trajectories: digestive, infectious, and mental conditions dominated trajectory origins in the Catalan network, whereas nervous system, neoplasm, and blood/immune conditions dominated in Denmark, with a unique Catalan enrichment for genitourinary-to-perinatal pathways (OR=2.19) absent from hospital data (Supplementary Figure 13; Supplementary Text).

## Discussion

Our primary finding is that the observation window length is a key determinant of which disease associations are detectable in primary care: 60.1% of significant comorbidity associations required at least 3 years of follow-up and were entirely invisible to studies using shorter windows. This systematic dependence is structured by disease category: Perinatal, Congenital, and Nervous system conditions showed the highest long-window dependence, while Ear, Musculoskeletal, and Respiratory disorders generated robust signals across all window lengths. The near-absence of early transient associations confirms that extending follow-up reveals previously undetected relationships rather than eliminating spurious ones. The moderate concordance of effect sizes with the Danish hospital network ^16^ (Spearman ρ=0.460) further supports the robustness of the primary care associations identified here, indicating that the core of the comorbidity structure is preserved across healthcare settings and populations.

To organise this temporal heterogeneity, we introduced the biological clock of multimorbidity: a two-dimensional framework that positions ICD-10 disease categories by their rates of comorbidity signal attenuation and the persistence of conditional risk. The four reproducible temporal patterns were robust under bootstrap resampling, leave-one-disease-out sensitivity analysis, and alternative clustering approaches, with Musculoskeletal, Genitourinary, Mental, and Pregnancy showing the highest positional stability (>95% quadrant retention). The transient-persistent pattern is best interpreted as a transitional zone between episodic and chronic progressive patterns rather than a discrete class, suggesting that the primary distinction in the clock space is between slow and fast cumulative decay, with persistent conditional risk as the more stable discriminating dimension. Within-category analysis revealed substantial heterogeneity (Endocrine/Metabolic and Mental disorders each spanned all four quadrants), indicating that chapter-level positions represent central tendencies that mask clinically relevant variation at finer diagnostic resolution. The total of 144,030 significant associations in the 0-5-year primary care network substantially exceeds the 62,821 reported by Jensen *et al* ^15^. in their foundational hospital-based cohort, but that study excluded pregnancy-related, symptom-based, injury, and administrative codes (categories representing a large fraction of primary care multimorbidity). Applying equivalent exclusions reduces our network to 87,026 associations, a figure that is 1.4-fold larger than the Jensen estimate and consistent with primary care capturing genuinely additional disease relationships that are invisible to hospital-based ascertainment ^33,34^.

The biological clock was systematically modulated by sex, particularly for Blood/Immune disorders, where women showed slower attenuation consistent with the predominance of chronic autoimmune conditions (notably lupus, rheumatoid arthritis, and Sjögren syndrome) whose comorbidities are expected to be more persistent than the acute haematological and reactive immune conditions more prevalent in men ^35,36^. Sex-based directional reversals were rare but concentrated in categories with established sexual dimorphism in immune regulation and inflammatory pathways, extending Westergaard *et al*.’s observation of reversed temporal directionality between sexes to a systematic, category-level characterisation ^16^. The temporal framework reveals a layer of sex-specific comorbidity structure that prevalence-based analyses alone do not capture: women showed higher persistent conditional risk across nearly all categories, and the asymmetric network overlap (with 85.2% of men’s associations present in the women’s network but only 61.4% of women’s associations present in men’s) is consistent with the higher multimorbidity burden documented in women in primary care, reflecting both higher healthcare utilisation and the broader spectrum of ambulatory conditions disproportionately affecting women ^37^.

Comparison with the Danish hospital network provides both external validation and contextual grounding for the primary care findings. The conserved backbone of shared associations, 2.7-fold more than expected by chance (hypergeometric test, p<10^-300^), with moderate but consistent concordance of effect sizes (Spearman ρ=0.460), confirms that the comorbidity structure identified in Catalan primary care reflects a genuine biological signal rather than a population-specific or setting-specific artefact. This validation is particularly relevant for the biological clock framework: the categories showing the strongest cross-system reproducibility (Ear, Blood/Immune, Mental, Circulatory) are precisely those with the most stable clock positions under bootstrap resampling, suggesting that temporally robust associations are also the most generalisable across healthcare settings. Beyond this validated core, the limited absolute overlap (3.6% of Catalan associations replicated in Denmark) is itself informative rather than a limitation. Primary care, by its nature, provides longitudinal follow-up of the general population across the full spectrum of disease severity, enabling detection of ambulatory, infectious, and ear/respiratory associations that would otherwise go undetected by hospital-based ascertainment. Hospital data, conversely, capture high-severity trajectories (Perinatal, Congenital, and Neoplasm transitions) that are under-represented in primary care. These two settings are therefore best understood as complementary lenses on the disease co-occurrence landscape, each revealing a segment of multimorbidity structure invisible to the other. The higher fold enrichment observed in sex-stratified analyses (women: 3.4-fold; men: 4.2-fold) further suggests that the conserved backbone is particularly robust for sex-specific disease relationships, reinforcing the sex-stratified findings reported above.

These findings have direct implications for the design and interpretation of EHR-based comorbidity studies and for clinical practice. Observation window length should be treated as a primary design parameter rather than a methodological detail, with selection informed by the temporal dynamics of the index condition: chronic progressive conditions require extended follow-up to capture their comorbidity burden, while episodic conditions are largely captured within the first year. These findings complement existing guidance on individualised multimorbidity management by providing an empirical basis for calibrating follow-up intervals to the temporal dynamics of the index condition, extending current frameworks to incorporate a quantitative, temporally resolved dimension that existing guidance lacks ^38,39^. Patients with index diseases in transient-persistent categories such as Circulatory, Digestive, and Symptoms disorders may harbour substantial undetected comorbidity risk that only becomes clinically manifest after several years: a study using a one-year observation window would miss 56-65% of the comorbidity associations detectable at five years for these categories, compared with only 15-25% for episodic categories such as Infectious and Pregnancy-related disorders. Conversely, for episodic categories, intensive short-term monitoring captures most of the comorbidity signal, and extended surveillance adds little clinical value. The near-absence of early transient associations (0.1%) confirms that comorbidities detected at short follow-up are almost universally persistent, reducing the risk that short-window screening generates transient false-positive associations. For patients with conditions in the chronic progressive quadrant (particularly musculoskeletal, ear, and respiratory disorders), the persistence of significant conditional risk across all annual intervals implies that the window for preventive intervention remains open throughout the observation period, not just in the immediate post-diagnosis phase. The complementarity between primary care and hospital networks further suggests that studies relying exclusively on hospital EHR data may systematically overlook ambulatory disease associations – primary care, by virtue of its longitudinal and population-wide coverage, is uniquely positioned to capture the full temporal arc of multimorbidity development.

The limited overlap between primary care and hospital comorbidity networks also has important implications for molecular comorbidity research. Studies of shared genetic architecture, transcriptomic similarity, and protein interaction networks routinely use EHR-derived comorbidity networks as ground truth to validate molecular hypotheses ^40^, but virtually all such networks have been derived from hospital registries. Our findings suggest that disease pairs with molecular evidence of shared biological mechanisms (potentially classified as false positives when benchmarked against hospital networks) may in fact represent true comorbidities detectable in primary care but absent from hospital data due to the ambulatory nature of the conditions involved. Incorporating primary care comorbidity networks as complementary ground truth, particularly for ambulatory, chronic, and symptom-based conditions, could substantially increase the sensitivity of molecular comorbidity studies and reduce the rate of false-negative molecular hypotheses. More broadly, the biological clock provides a temporal dimension currently absent from molecular analyses of comorbidity: disease pairs in the chronic progressive quadrant, where conditional risk persists throughout the observation period, are stronger candidates for shared biological mechanisms than episodic pairs whose co-occurrence is concentrated in the acute phase ^41^.

## Limitations

Several limitations warrant consideration. First, the closed cohort design excludes individuals who entered the healthcare system after January 2008, limiting representativeness for paediatric conditions and recently arrived populations; left-truncation may additionally inflate short-window associations for conditions prevalent at baseline, though longer-window associations are less susceptible to this bias. The matched design additionally does not account for factors such as socioeconomic status, lifestyle, or treatment patterns, which may influence the observed comorbidity structure; in particular, socioeconomic deprivation is a well-established driver of multimorbidity clustering and may account for a subset of the observed co-occurrences ^1,42,43^. Second, the analysis is restricted to primary care diagnoses; conditions managed exclusively in hospital or specialist settings may be under-captured, potentially leading to underestimation of comorbidities for severe or acute conditions that bypass primary care. Third, ICD-10 coding in primary care may be influenced by help-seeking behaviour and diagnostic labelling practices, particularly for symptom-based codes. While this is not unique to primary care, it may be more pronounced in ambulatory settings, where diagnostic certainty is lower than in hospital records. Fourth, the comparison with Danish hospital data is cross-system rather than within-system; the two datasets differ in follow-up duration, coding practices, and population composition, and the Jaccard index and Spearman correlation should be interpreted as lower bounds on the true underlying comorbidity structure rather than definitive measures of cross-system reproducibility. Fifth, the requirement for five years of follow-up after index diagnosis introduces survivorship bias, as individuals who died or were lost to follow-up within five years are excluded; reported associations may therefore underestimate comorbidity risk in more severe or rapidly progressing disease presentations. Sixth, the biological clock was internally robust to resampling and sensitivity analyses but could not be externally validated on an independent dataset with comparable temporal resolution, because the Danish registry data were available only as aggregated disease trajectories, lacking the nine-window structure required for direct replication. The choice of aggregation function underlying the ultrametric closure also affects which comorbidity pathways are recovered, though not the set of reachable disease pairs; alternative aggregation functions are discussed in the appendix. Finally, this study characterises comorbidity at the population level through directed disease-pair associations; this approach is complementary to but distinct from person-centred analyses, which focus on the number and combination of conditions within individual patients and their clinical trajectories over time.

## Conclusion

The temporal structure of multimorbidity is not captured by any single observation window: most disease associations require extended follow-up to become detectable, and the rate at which they emerge varies systematically across disease categories. Although demonstrated here in primary care (where longitudinal population-wide follow-up makes this quantification possible), the implications extend to any EHR-based comorbidity study in which observation window length is treated as a methodological detail rather than a primary design parameter. The biological clock of multimorbidity provides a framework for understanding how disease associations emerge and persist over time, with direct consequences for downstream analyses: as demonstrated here for sex-stratified comorbidity dynamics, the temporal dimension reveals structure that static or short-window approaches fail to capture. The complementarity of primary care and hospital networks further underscores the need to view these settings as distinct yet synergistic sources of knowledge on multimorbidity.

## Methods

### Study population and data sources

Catalonia is an autonomous community of Spain with a population of approximately 7.5 million inhabitants. We analysed 11 years of anonymised electronic health record (EHR) data, spanning 1 January 2008 – 31 December 2018, provided by the Catalan Health Institute (CHI) and extracted from SIDIAP (Information System for Research in Primary Care). The CHI manages primary healthcare teams serving approximately 74% of the Catalan population. Since 2005, all CHI healthcare professionals have used the same electronic clinical record system (e-CAP) to register demographic information, prescriptions, laboratory tests and diagnoses during routine medical and nursing visits. These data were originally collected for clinical and administrative purposes. Individuals were eligible if they were aged over 1 year on 1 January 2008 and had at least one recorded visit during the study period. Follow-up was censored at death or transfer to another primary health care team.

Diagnoses were coded according to the International Classification of Diseases, 10th revision (ICD-10), with recording available up to the five-digit level. For the present study, diagnoses were aggregated at the three-digit ICD-10 level. Only individuals born before January 2007 were included to ensure complete diagnostic capture during the observation window.

The study cohort comprised 5,821,197 individuals with at least one recorded diagnosis during the study period, including 2,989,599 women (51.4%) and 2,831,598 men (48.6%). The cohort was closed at baseline in January 2008, such that only individuals already registered in the system at that time were followed longitudinally through December 2018. New entrants after 2008 were not included. Consequently, the cohort reflects a fixed baseline population and does not incorporate younger incident birth cohorts during follow-up.

The study was approved by the ethics committee of the Jordi Gol University Institute for Research in Primary Healthcare. This manuscript was not prepared in collaboration with the registry, and the interpretations and conclusions expressed herein are solely those of the authors.

### Temporal disease co-occurrence analysis

To characterise temporal relationships between diseases, we evaluated co-occurrence patterns across all ordered pairs of ICD-10 three-digit diagnoses. Only the first recorded occurrence of each diagnosis was retained per individual. For each disease A with at least 100 patients and a period prevalence not exceeding 20%, patients with a first recorded diagnosis of A during the study period were defined as the exposed cohort, and the date of that diagnosis was used as the index date. The 20% prevalence threshold was applied because higher prevalences leave an insufficient pool of unexposed individuals to identify five matched controls per exposed patient; this criterion resulted in the exclusion of three diseases (M54, J00, and T14). For each exposed patient, five matched unexposed individuals were randomly selected from individuals without a diagnosis of disease A at any point during the study period. Exposed patients were required to have received at least one other diagnosis within ±2 months of the exposed patients’ index date and were matched on sex and five-year age group. This matching scheme was designed to reduce confounding due to demographic differences and differential healthcare utilisation. Exposed and unexposed individuals were required to have at least 5 years of follow-up after the index date to ensure that the full 0-5-year observation window was available for all individuals. The same matched cohorts were used across all temporal analyses for a given disease A; only the observation window applied to detect subsequent diagnoses varied between analyses. Analyses were performed separately in women, men, and the combined population.

### Cumulative and conditional follow-up windows

For each ordered disease pair A → B, we evaluated whether individuals diagnosed with disease A had an increased probability of subsequently receiving a diagnosis of disease B compared with matched unexposed individuals, using two complementary sets of follow-up windows.

Cumulative windows spanned from the index date to a fixed endpoint after diagnosis of A: 0-1, 0-2, 0-3, 0-4, and 0-5 years. These windows quantify the overall probability of developing B over a given period, regardless of when B first occurs during that period. Conditional windows spanned non-overlapping successive intervals: 1-2, 2-3, 3-4, and 4-5 years after the index date. Each conditional window quantifies the risk of developing B within a specific interval, given that B had not been diagnosed in any earlier window. Individuals who developed B before the start of a given conditional window were excluded from the analysis for that window. These windows directly estimate the residual or late-emerging risk of B in patients who have not yet developed the secondary condition, complementing the cumulative analysis. These conditional probabilities correspond mathematically to discrete-time hazards, as they quantify the probability of developing the outcome during a given interval conditional on remaining event-free until the start of that interval. However, we use the term conditional risk throughout to avoid confusion with continuous-time hazard models.

For each disease pair and time window, we constructed a 2×2 contingency table comparing the occurrence of disease B among the exposed cohort and comparison group: a, individuals with disease A who developed disease B within the window; b, individuals with disease A who did not develop disease B; c, unexposed individuals who developed disease B; and d, unexposed individuals who did not. A continuity correction of 0.5 was added to all cells to avoid instability due to zero counts. From these counts, we computed log relative risks using Morris and Gardner’s formula ^44^ and their standard errors.

Disease pairs involving ICD-10 dagger–asterisk combinations were identified and removed. Dagger-asterisk pairs represent aetiology–manifestation coding relationships rather than independent disease co-occurrences; their inclusion would artificially inflate the comorbidity network with coding artefacts. A total of 1,873 three-digit dagger-asterisk pairs were identified and excluded.

### Empirical Bayes shrinkage of relative risk estimates

The large number of disease pairs evaluated and the wide range of disease prevalences can yield unstable relative risk estimates, particularly for pairs involving rare conditions or small numbers of exposed individuals. To address this, we applied empirical Bayes shrinkage using the adaptive shrinkage framework ^45^. Observed log relative risks were modelled as noisy estimates of underlying true effect sizes, with the prior distribution estimated empirically as a data-driven mixture of zero-centred normal distributions. This approach adaptively shrinks unstable estimates towards zero while preserving well-supported large effects.

From the fitted posterior distribution, we extracted for each disease pair the posterior mean log relative risk, the posterior standard deviation, and the local false sign rate (LFSR), defined as the posterior probability that the true direction of effect is opposite to the observed estimate. Disease pairs were classified as robust temporal co-occurrence associations if they satisfied three criteria simultaneously: LFSR ≤ 0.05, a lower bound of the 95% credible interval for the posterior relative risk ≥ 1.01, and at least 100 individuals diagnosed with both diseases within the observation window. Only associations satisfying all three criteria were retained for network construction and downstream analyses. These filtered associations were used to construct temporal comorbidity networks for each time window and sex stratum.

### Assessment of comorbidity directionality

For each unordered disease pair {A, B}, directionality was assessed by comparing the number of observed transitions A → B with the number of reverse transitions B → A, following the approach of Jensen *et al.* ^15^. For each directed pair A → B, the directionality parameter θ(A → B) was defined as the proportion of transitions in the preferred direction:

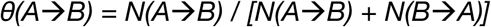

where N(A → B) denotes the number of exposed individuals with disease A who developed disease B, and N(B → A) denotes the corresponding count from the reverse analysis. A one-sided binomial test was applied to evaluate whether A → B occurred more frequently than expected under a symmetric null (p = 0.5). P values were adjusted within each window and population stratum using the Benjamini-Hochberg procedure. A pair was classified as having a preferred direction A → B if the FDR-adjusted p-value was ≤ 0.05 and θ > 0.5.

### The biological clock of multimorbidity: temporal detectability and dynamics

To characterise the temporal dynamics of comorbidity associations, we analysed the trajectory of the posterior relative risk across the five cumulative windows for all disease pairs that were significant in at least one window (persistent pairs). For each persistent pair, a weighted linear regression of the posterior RR on follow-up time (years 1-5) was fitted, with observations weighted by the inverse squared posterior standard error. The slope coefficient quantifies the rate of change in comorbidity signal detectability over time (hereafter referred to as the attenuation gradient) with negative values indicating weakening of the association as follow-up extends, and positive values indicating strengthening.

Category-level attenuation gradients were summarised as the median weighted slope across all persistent pairs within each ICD-10 category of the index disease. Heterogeneity across categories was assessed using a Kruskal-Wallis test, and post hoc pairwise comparisons between each category and the most stable reference category (Endocrine/Metabolic disorders) were performed using Wilcoxon rank-sum tests with a Benjamini-Hochberg correction.

To characterise residual conditional risk, each disease pair with data in all four conditional windows was classified according to its pattern of conditional significance, using a significance threshold of LFSR < 0.05, lower bound of the 95% credible interval ≥ 1.01, and ≥ 100 exposed individuals. Pairs were classified as showing persistent conditional risk if they were significant in at least one early conditional window (1-2 or 2-3 years) and in at least one late conditional window (3-4 or 4-5 years); as showing late-emerging conditional risk if significant only in the late conditional windows (3-4 or 4-5 years) but not in either early window; as showing early-only risk if significant only in the early windows (1-2 or 2-3 years) without late conditional signal; and as not significant otherwise. This classification used all four conditional windows symmetrically, including the 2-3-year interval, to avoid artificially excluding transitional risk that emerges between the first and third years after diagnosis.

The two temporal dimensions – rate of cumulative attenuation (median-weighted slope) and the proportion of pairs with persistent conditional risk – were combined to define a two-dimensional biological clock space. The denominator for the proportion of pairs with persistent conditional risk comprised all disease pairs with data in all four conditional windows, irrespective of their significance in the cumulative windows. Quadrant boundaries were set at the median values of both dimensions across all ICD-10 categories, yielding a data-driven classification. The four resulting quadrants were interpreted as: episodic (fast decay, low conditional persistence), chronic stable (slow decay, low conditional persistence), chronic progressive (slow decay, high conditional persistence), and transient-persistent (fast decay, high conditional persistence). The transient-persistent pattern is a category-level property reflecting the combination of fast cumulative decay and a high proportion of transient-persistent conditional risk across the category; individual pairs within transient-persistent categories may show transient-persistent rather than strictly transient-persistent conditional trajectories.

Sex-stratified biological clock analyses were performed by repeating all slope and conditional pattern calculations independently for women and men. Directional reversals between sexes were identified as disease pairs for which A → B was the preferred direction in women but B → A was the preferred direction in men in the 0-5-year network, using the binomial test approach described above. Directional reversals across temporal windows were identified as pairs in which A → B was preferred in the 0-1 year network but B → A was preferred in the 0-5 year network.

### Robustness analysis of the biological clock

The stability of ICD-10 category positions in the two-dimensional biological clock space was assessed through three complementary analyses. First, a pair-level bootstrap was performed by resampling disease pairs with replacement within each ICD-10 chapter (1,000 replicates, stratified by chapter to preserve category sample sizes). In each replicate, the median weighted cumulative slope and proportion of pairs with persistent conditional risk were recomputed for each category, quadrant boundaries were recalculated as the medians of both dimensions across all categories, and each category was assigned to a quadrant. Quadrant retention was defined as the proportion of bootstrap replicates in which a category retained its original quadrant assignment. Second, a leave-one-index-disease-out sensitivity analysis was performed by iteratively removing all persistent comorbidity pairs originating from each index disease within a category and recomputing the category’s clock position. This addressed the potential influence of highly connected individual diseases on the chapter-level summary. Third, the sensitivity of quadrant assignments to the boundary definition method was assessed by comparing the median-split classification with a k-means clustering (k=4) applied to normalised clock coordinates, using 50 random initialisations to ensure convergence.

### Temporal detection taxonomy

To characterise the complete temporal signature of each disease pair, all nine windows (five cumulative and four conditional) were combined into a single classification. Each pair was assigned to one of nine mutually exclusive classes based on the combination of windows in which it was detected as significant: omnipresent (significant in all nine windows); persistent cumulative with partial conditional (significant in all five cumulative windows and at least one conditional window); persistent cumulative only (significant in all five cumulative windows but no conditional windows); long-window only (significant only in cumulative windows of three or more years, with no conditional signal); late cumulative with conditional risk (significant only in longer cumulative windows with additional conditional signal); conditional only (significant exclusively in conditional windows); early transient (significant only in the 0-1 year window); early cumulative with late conditional (significant in early cumulative windows with late conditional signal); and mixed or partial (all remaining patterns).

The distribution of temporal classes across ICD-10 categories was summarised as the proportion of pairs in each class within each category, separately for index and secondary disease categories, and for category pairs.

### Prospective comorbidities from graph closure

The comorbidity network was represented as a directed weighted graph G = (V, E, w), with edge weights w_(i,j)_ quantifying the relative risk, RR_(i,j)_, of disease j following i. To consider positive comorbidity associations, we set w_(i,j)_ = RR_(i,j)_ if RR_(i,j)_ > 1, otherwise RR_(i,j)_ = 0, i.e. the absence of a positive association. The graph ultrametric closure G^t,u^ (a generalisation of graph reachability to weighted graphs, considering a prospective relative risk determined by the lowest relative risk within a disease trajectory) was computed over G, yielding a graph in whose non-zero edge weights indicate the existence of a comorbidity pathway (direct or indirect) between diseases i and j ^46,47^. When comparing two graphs, G_1_ and G_2_, the ultrametric closure was computed independently for each and restricted to shared diseases; the odds ratio (OR) of prospective comorbidity between disease groups was computed to quantify differential graph closure.

### Sex-biased disease prevalence and category enrichment analysis

Sex differences in disease prevalence were assessed using aggregated counts of individuals with at least one recorded diagnosis between 2008 and 2018, stratified by sex. For each ICD-10 three-digit disease, we constructed a 2×2 contingency table contrasting sex (women versus men) and disease status (diagnosed versus not diagnosed), using the total number of women and men under observation as denominators. Because diagnostic data prior to 2008 were unavailable, these estimates represent recorded period prevalence within the study window.

Differences in prevalence were evaluated using two-sided Fisher’s exact tests. Effect sizes were quantified as odds ratios (ORs), defined as the odds of disease in women relative to men. P values were adjusted across diseases using the Benjamini-Hochberg false discovery rate (FDR) procedure. Diseases were classified as sex-biased if they met both statistical and effect-size criteria (FDR ≤ 0.05 and OR ≥ 1.3 for higher prevalence in women, or OR ≤ 1/1.3 for higher prevalence in men).

To determine whether sex-biased diseases were concentrated within particular ICD-10 chapters, enrichment analyses were performed separately for women- and men-biased sets using two-sided Fisher’s exact tests on 2×2 contingency tables (in-category versus out-of-category; biased versus non-biased). P values were adjusted across categories using the Benjamini-Hochberg procedure.

### Statistical analysis of sex differences in age at first diagnosis

For each disease, we extracted the age at first recorded diagnosis per individual. The mean and standard deviation of age at first diagnosis were calculated separately for women and men. Sex differences were assessed using two-sided Welch’s t-tests, which do not assume equal variances. P values were adjusted across diseases using the Benjamini-Hochberg FDR procedure.

For each disease, the mean age difference was defined as the mean age in women minus the mean age in men, such that positive values indicate later diagnosis in women and negative values indicate later diagnosis in men. To evaluate whether specific ICD-10 categories exhibited systematic differences in diagnostic timing, we analysed disease-specific mean age differences using inverse-variance weighted linear models. For each disease, the standard error of the mean difference was calculated from the sex-specific standard deviations and sample sizes. Category-level effects were summarised as average age differences in years with corresponding 95% confidence intervals. P values were adjusted across categories using the Benjamini-Hochberg procedure. As a sensitivity analysis, we additionally restricted the analysis to diseases showing significant sex differences and tested whether the proportion of diseases diagnosed later in women within each category differed from 0.5 using one-sided binomial tests, again adjusting for multiple comparisons across categories.

### Cross-population comparison with Danish hospital data

To assess the reproducibility of primary care comorbidity networks across healthcare systems, we compared the Catalan 0-5-year networks with the hospital-based Danish disease trajectory data reported by Westergaard *et al*. ^16^. Both studies applied a matched cohort design with 1:5 matching, three-digit ICD-10 disease codes, and the same minimum event threshold (≥100 patients per pair), filtering associations to a lower confidence bound for the relative risk ≥ 1.01. The two studies differed in healthcare setting (primary care versus hospital admissions), follow-up window (a fixed 0-5 year window in the present study versus complete registry follow-up across a 21-year period in Westergaard *et al*. ^16^), matching criteria (sex, five-year age group, and date of index diagnosis ±2 months in the present study versus five-year age group, type of hospital encounter, and discharge month and year ±3 months in Westergaard *et al*. ^16^), and shrinkage approach (empirical Bayes via adaptive shrinkage in the present study versus fully Bayesian hierarchical modelling via Hamiltonian Monte Carlo sampling in Westergaard *et al*. ^16^). Cross-population analyses were restricted to the 1,204 ICD-10 three-digit diseases present in the studyable universe of both datasets – defined as diseases appearing in at least one significant or tested pair in each dataset, irrespective of whether they achieved significance – yielding a common disease universe for all comparisons.

Network overlap between the two populations was quantified using three complementary metrics: the Jaccard index (number of shared pairs divided by the total number of pairs), the proportion of Catalan pairs in the Danish network, and the proportion of Danish pairs in the Catalan network. These metrics were computed separately for the combined population, women, and men.

For disease pairs significant in both datasets, concordance of effect size was assessed using Spearman’s rank correlation between log-scale relative risks. Correlations were computed for the combined population and sex-stratified analyses. Category-pair-specific concordance was assessed by computing Spearman correlations within each ICD-10 category combination, restricted to pairs with at least 10 shared observations. Enrichment of specific ICD-10 category transitions among shared pairs relative to the union of all significant associations in either dataset was assessed using Fisher’s exact tests on a 2×2 contingency table cross-classifying each pair by presence in Catalonia (yes/no) and presence in Denmark (yes/no). This formulation accounts for pairs present in Denmark but absent from Catalonia, providing a symmetric assessment of enrichment relative to the full cross-system universe. Separately, differential representation of category transitions between the two healthcare systems was assessed by comparing the proportion of each category combination among all Catalan significant pairs against its proportion among all Danish significant pairs, using Fisher’s exact tests. All p-values were adjusted using the Benjamini-Hochberg procedure.

Directional concordance between the two healthcare systems was assessed for shared directed pairs – pairs for which a preferred direction was identified in both datasets. A pair was classified as directionally concordant if the same direction (A → B) was preferred in both Catalonia and Denmark. Directional reversals between healthcare systems were identified as pairs in which A → B was preferred in Catalonia but B → A was preferred in Denmark, using the same binomial test-based definition of preferred direction described above and applied independently to the Danish data.

### Network centrality analysis: identification of multimorbidity initiators and sinks

To characterise the structural role of individual diseases in the propagation of multimorbidity beyond direct pairwise associations, we represented the 0-5-year combined-population comorbidity network as a weighted directed graph, in which nodes correspond to three-digit ICD-10 diseases and directed edges represent significant comorbidity associations. Three weighting schemes were applied to edges: the posterior shrunk relative risk, the number of incident secondary cases, and their product, yielding three parallel network representations with identical topology but differing edge weights. Prior to analysis, three diseases with period prevalence exceeding 20% (M54, J00, T14) were excluded to avoid hub artefacts driven by highly prevalent conditions that would dominate centrality metrics irrespective of their specific comorbidity structure. Diseases that generated no significant outgoing associations (out-dangling nodes) were likewise excluded.

For each disease and each weighting scheme, two centrality metrics were computed. Weighted outdegree was defined as the sum of edge weights across all outgoing associations from a given disease, reflecting the total direct comorbidity burden generated by that condition in terms of both the number and magnitude of its associations. PageRank ^48^ was computed using the standard random-walk formulation with a damping factor of 0.85, applied to the weighted directed graph, reflecting the importance of a disease as a destination weighted by the centrality of its source diseases – that is, a disease receives high PageRank if it is the target of associations originating from other high-PageRank diseases. Both metrics were converted to ranks (rank 0 = highest value) to enable direct comparison across weighting schemes.

For each disease and weighting scheme, a discrepancy score was defined as the difference between the PageRank rank and the weighted outdegree rank. Positive discrepancy indicates that a disease ranks higher by weighted outdegree than by PageRank – consistent with an initiator profile, in which a condition generates broad and high-magnitude direct associations whose downstream targets are not themselves network hubs. Negative discrepancy indicates that a disease ranks higher by PageRank than by weighted outdegree – consistent with a sink profile, in which a condition occupies a central network position as the convergent destination of multiple upstream trajectories despite generating comparatively few or lower-magnitude direct associations.

Discrepancy scores were standardised to z-scores within each weighting scheme (mean = 0, SD = 1). A disease was classified as a robust initiator if its z-score exceeded +2 in at least two of the three weighting schemes, and as a robust sink if its z-score fell below −2 in at least two of the three weighting schemes. This threshold corresponds approximately to the top and bottom 2.5% of the discrepancy distribution under each weighting scheme, and the requirement for consistency across at least two weighting schemes reduces the influence of artefacts specific to any single edge weight definition.

## Author contributions

J.S.-V., A.V., C.Z., and F.X.C. designed all experiments. J.S.-V., C.Z., and F.X.C. performed the experiments. A.N.-M., D.C., L.M.R., and C.V. provided technical advice. J.S.-V. wrote the manuscript. All authors discussed the results and commented on the manuscript.

## Data Availability

The primary care data analyzed are only accessible upon request and following approval of an analysis protocol by both the Scientific Committee and the Ethics Committee of IDIAP Jordi Gol. The results generated from these data are presented in the manuscript.

