## Supplementary Material for "The biological clock of multimorbidity: temporal dynamics of disease co-occurrence in primary care"

### Supplementary Information

#### *Supplementary Text 1. Cross-cohort concordance of sex-biased disease prevalence patterns*

At the category level, several enrichment patterns were reproducible across cohorts. Endocrine, nutritional and metabolic diseases were significantly enriched among women-biased conditions in both Catalonia and Denmark, as were skin and musculoskeletal disorders, suggesting a stable women predominance in chronic metabolic, dermatological and connective tissue diseases across care settings. Similarly, men predominance in neoplastic diseases was observed in both populations, consistent with known sex disparities in cancer incidence.

However, notable divergences emerged in specific domains. In Denmark, injury and poisoning showed strong men enrichment, whereas in Catalonia the pattern was more heterogeneous. Mental and behavioural disorders were enriched among men-biased diseases in Catalonia but among women-biased diseases in Denmark, highlighting potential differences in case ascertainment between primary and hospital care. Respiratory diseases were significantly underrepresented among women-biased diseases in Denmark, contrasting with the clearer male enrichment observed in Catalonia.

Disease-level discordances further illustrated these population-specific effects. For example, chronic hepatitis (B18) was more prevalent in women in Denmark but in men in Catalonia (Supplementary Table 6). Similarly, malignant neoplasm of the anus and anal canal was women-biased in Denmark but men-biased in Catalonia. Conversely, certain trauma-related conditions, including burns of the wrist and hand, were men-biased in Denmark but women-biased in Catalonia. These reversals were largely concentrated in infectious, injury-related and selected neoplastic categories. Together, these findings indicate that while many sex-biased prevalence patterns are consistent across populations and healthcare levels, a meaningful subset of diseases exhibit setting-specific directionality.

#### *Supplementary Text 2. Cross-cohort concordance of sex differences in age at diagnosis*

In a sensitivity analysis restricted to diseases with significant sex differences at the individual-disease level, directional enrichment tests yielded consistent results. Categories such as Injury and poisoning, Circulatory system, Musculoskeletal system and connective tissue, and Mental and behavioural disorders showed a significant excess of diseases diagnosed later in women. No category showed a significant excess of later-diagnosed diseases in men after correction for multiple testing.

To assess the robustness and generalizability of sex differences in diagnostic timing, we compared disease-specific mean age differences (women – men) between Catalonia and Denmark. Overall, the direction of effects was largely concordant across populations, with most diseases clustering along the positive diagonal in the cross-cohort comparison, indicating similar sex-specific shifts in age at diagnosis (see Supplementary Figure 14).

The overlap analysis showed that 605 diseases exhibited significant sex differences in both populations (512 later in women, and 93 in men), supporting shared biological and/or health-system–independent components of sex-differential diagnostic timing.

On the other hand, 282 were significant only in Denmark and 151 only in Catalonia (see Supplementary Figure 14).

Representative examples of cross-cohort agreement include type 2 diabetes, for which women were diagnosed later in both Denmark (2.68 years) and Catalonia (4.78 years), and dementia in Alzheimer's disease, also diagnosed later in women in both settings (2.1 and 1.66 years, respectively). Similarly, several circulatory and metabolic disorders showed consistent women delays across cohorts.

In contrast, a subset of diseases showed opposing effects. For example, certain infectious diseases (such as A17) showed a later diagnosis in women in Catalonia but in men in Denmark. Endocrine disorders such as E16 also displayed cross-cohort divergence, with opposite signs of mean age difference despite statistical significance in both populations. These discordant patterns likely reflect differences in case ascertainment, disease severity thresholds or healthcare utilisation pathways.

Together, these analyses indicate that while the predominant pattern of sex differences in age at diagnosis is reproducible across healthcare systems, a non-negligible fraction of diseases exhibit context-dependent reversals in directionality, underscoring the complementary information captured by primary and secondary care data.

***Supplementary Text 3. Network reachability across temporal windows***

Comorbidity associations can be modelled as a directed graph, where nodes represent diseases and edges represent significant directed associations. Beyond direct associations, this representation allows characterisation of multi-step disease trajectories through graph reachability – the set of diseases that can eventually be reached from a given index disease through one or more intermediate conditions.

The 144,030 directed associations in the 0-5-year network imply 643,224 eventual comorbidities among the same disease universe, compared with 354,728 eventual comorbidities implied by the 56,084 associations in the 0–1-year network. This 1.8-fold increase in eventual comorbidities, relative to the 1.8-fold increase in direct associations, indicates that longer follow-up windows expand the reachable disease landscape proportionally but with qualitatively different trajectory structures.

Among disease categories enriched as trajectory origins, the 1-year window showed disproportionate enrichment for digestive (OR = 1.23) and respiratory (OR = 1.15) index conditions, consistent with these categories generating dense local clusters of short-latency associations. The 5-year window showed stronger enrichment for neoplasms (OR = 1.17) and congenital malformations (OR = 3.16) as trajectory origins, consistent with the long latency of oncological sequelae and developmental consequences identified in the temporal detection taxonomy. Both windows converged on similar destination categories – digestive, genitourinary, musculoskeletal, and symptom-based conditions – indicating that the qualitative differences between windows are primarily in trajectory origins rather than destinations (Supplementary Figure 10).

***Supplementary Text 4. Network reachability comparison between Catalan and Danish networks***

To assess whether the complementarity between primary care and hospital comorbidity networks extends beyond direct associations to multi-step disease trajectories, we compared the graph reachability of the Catalan 0–5-year and Danish networks restricted to the 1,204 shared diseases.

Despite limited structural overlap in direct associations (Jaccard = 0.033), the two networks showed convergent destination categories – shared diseases in both populations eventually led to conditions corresponding to the same broad ICD-10 categories. However, trajectory origins differed systematically between systems. In the Catalan network, trajectory origins were disproportionately enriched for digestive (OR = 1.29), infectious and parasitic (OR = 1.12), and mental and behavioural (OR = 1.15) conditions, consistent with the ambulatory profile of primary care. In the Danish hospital network, trajectory origins were enriched for nervous system (OR = 1.35), neoplasms (OR = 1.51), and blood and blood-forming organs (OR = 1.60) conditions, consistent with the high-severity profile of hospital-ascertained multimorbidity.

A unique feature of the Catalan network was the enrichment of genitourinary conditions as trajectory origins leading to perinatal conditions (OR = 2.19), a pathway entirely absent from the Danish hospital network. This is consistent with the predominance of obstetric and reproductive trajectories in primary care – conditions that rarely lead to hospital admissions but represent clinically important multi-step pathways from genitourinary morbidity to perinatal outcomes, captured exclusively in ambulatory records.

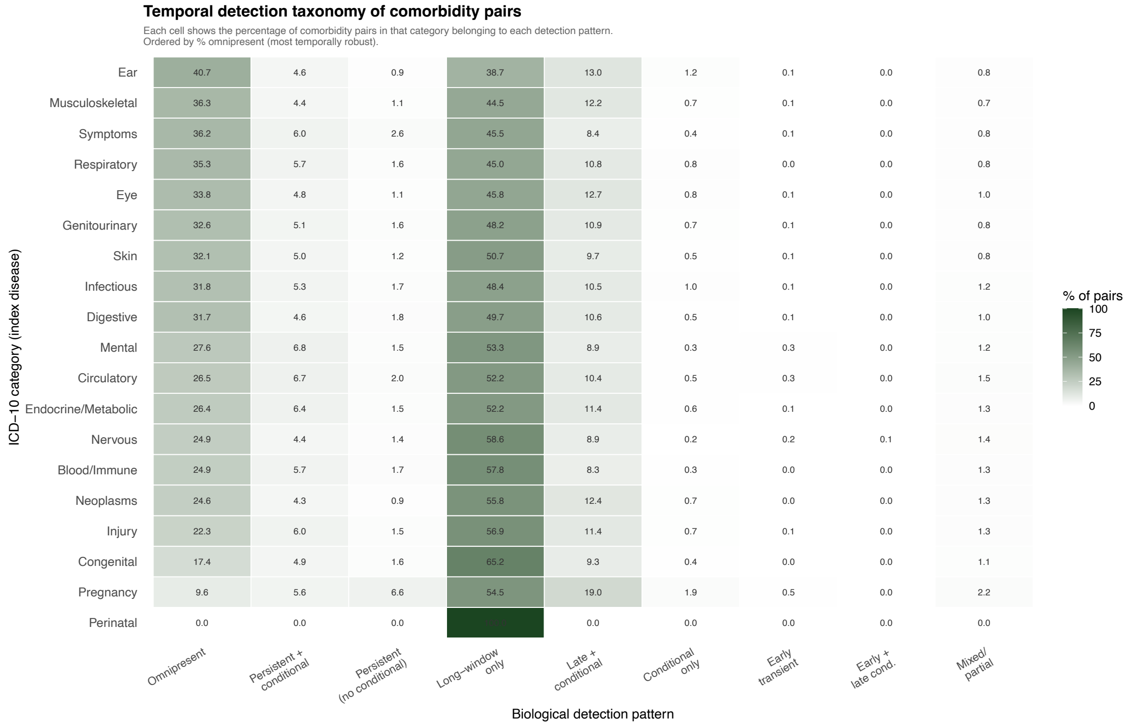


**Supplementary Figure 1**. Detailed temporal detection taxonomy of comorbidity pairs by ICD-10 category. Each cell shows the percentage of comorbidity pairs in that category (index disease, rows) belonging to each of nine temporal detection patterns (columns), defined by the combination of cumulative (0-1 to 0-5 years) and conditional (1-2 to 4-5 years) windows in which the association is significant (LFSR < 0.05, CI lower bound ≥ 1.01, ≥100 exposed individuals). Detection patterns are ordered from left to right by temporal robustness: Omnipresent = detectable in all nine windows; Persistent + conditional = detectable in all five cumulative windows with at least one significant conditional window; Persistent (no conditional) = detectable in all five cumulative windows without conditional signal; Long-window only = detectable only in cumulative windows of three or more years without any conditional signal; Late + conditional = detectable in longer cumulative windows with additional conditional risk; Conditional only = detectable exclusively in conditional windows; Early transient = detectable only in the 0-1 year window; Early + late conditional = detectable in early cumulative windows with late conditional signal; Mixed/partial = all remaining patterns. Categories are ordered by decreasing proportion of omnipresent pairs. Colour intensity reflects the percentage of pairs (white = 0%, dark green = 100%). Values ≥ 5% are shown numerically. Analysis performed for the combined population (both sexes).

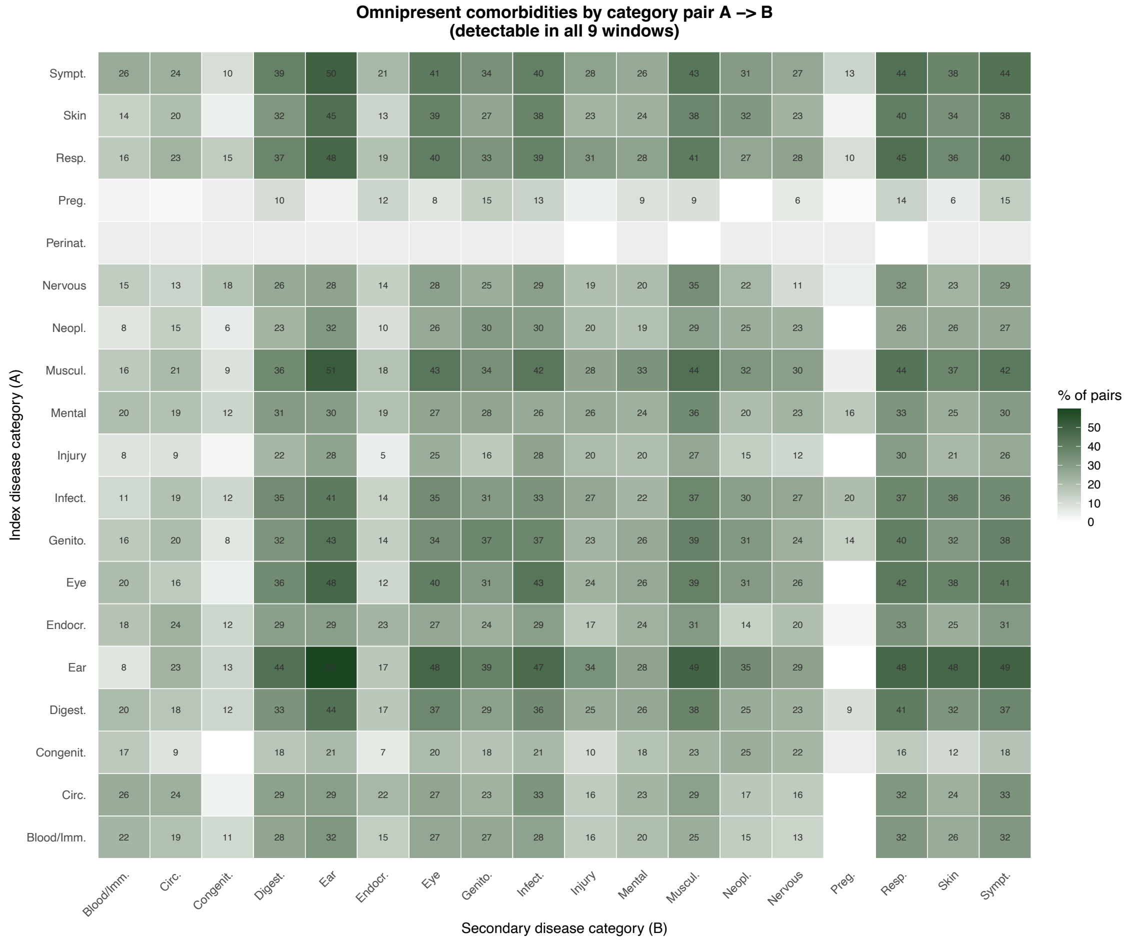


**Supplementary Figure 2.** Distribution of omnipresent comorbidity pairs (detectable in all nine follow-up windows) across ICD-10 category pairs. Each cell shows the percentage of pairs in that category combination classified as omnipresent. Categories are ordered by decreasing mean omnipresent percentage. Numbers shown for cells ≥ 5%. Ear disorders as secondary disease show consistently high omnipresent proportions across index categories, reflecting the chronic and recurrent nature of auditory comorbidities.

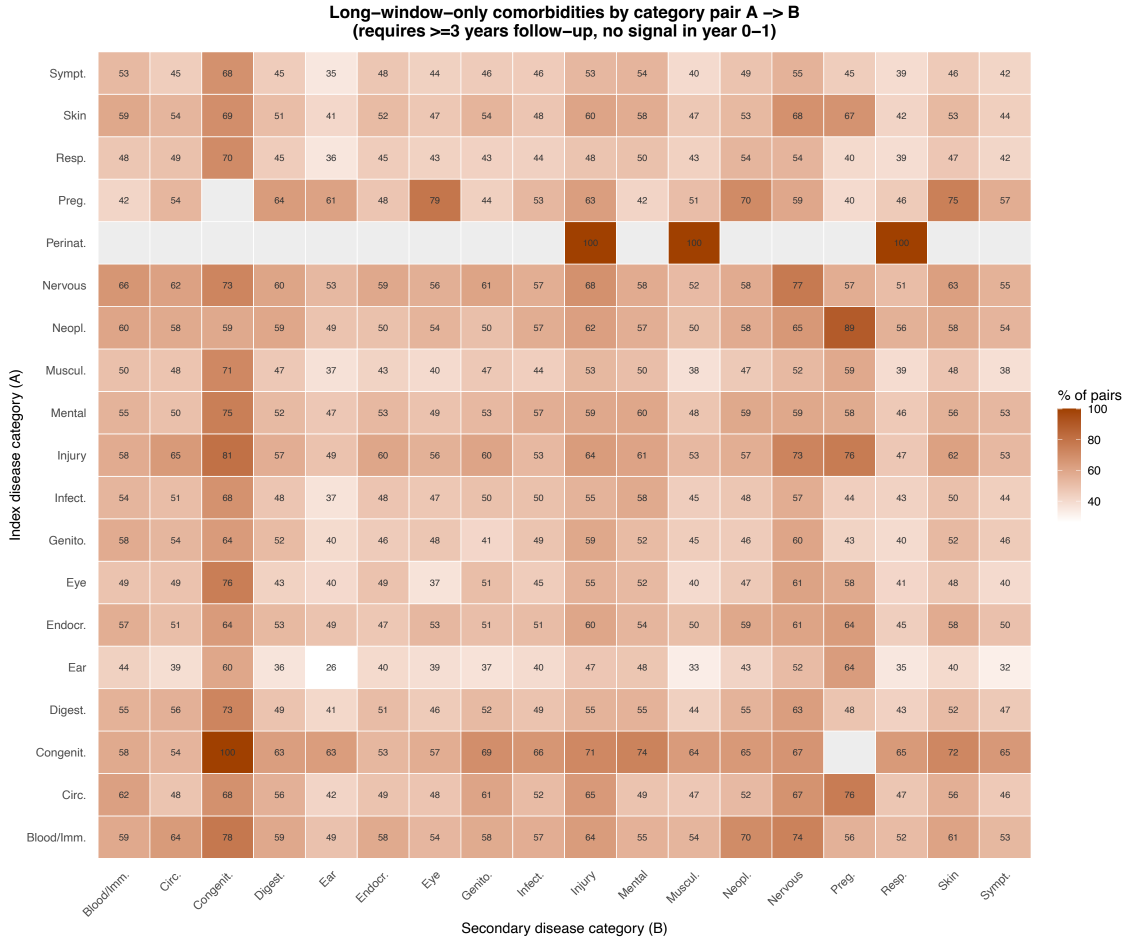


**Supplementary Figure 3.** Distribution of long-window-only comorbidity pairs (detectable only in cumulative windows of ≥3 years, with no signal in the 0-1 year window and no conditional risk) across ICD-10 category pairs. Perinatal conditions as index disease show 100% long-window-only pairs across all secondary categories. Congenital → Congenital and Neoplasms → Pregnancy also show very high proportions (100% and 89% respectively), reflecting the long latency of these disease relationships.

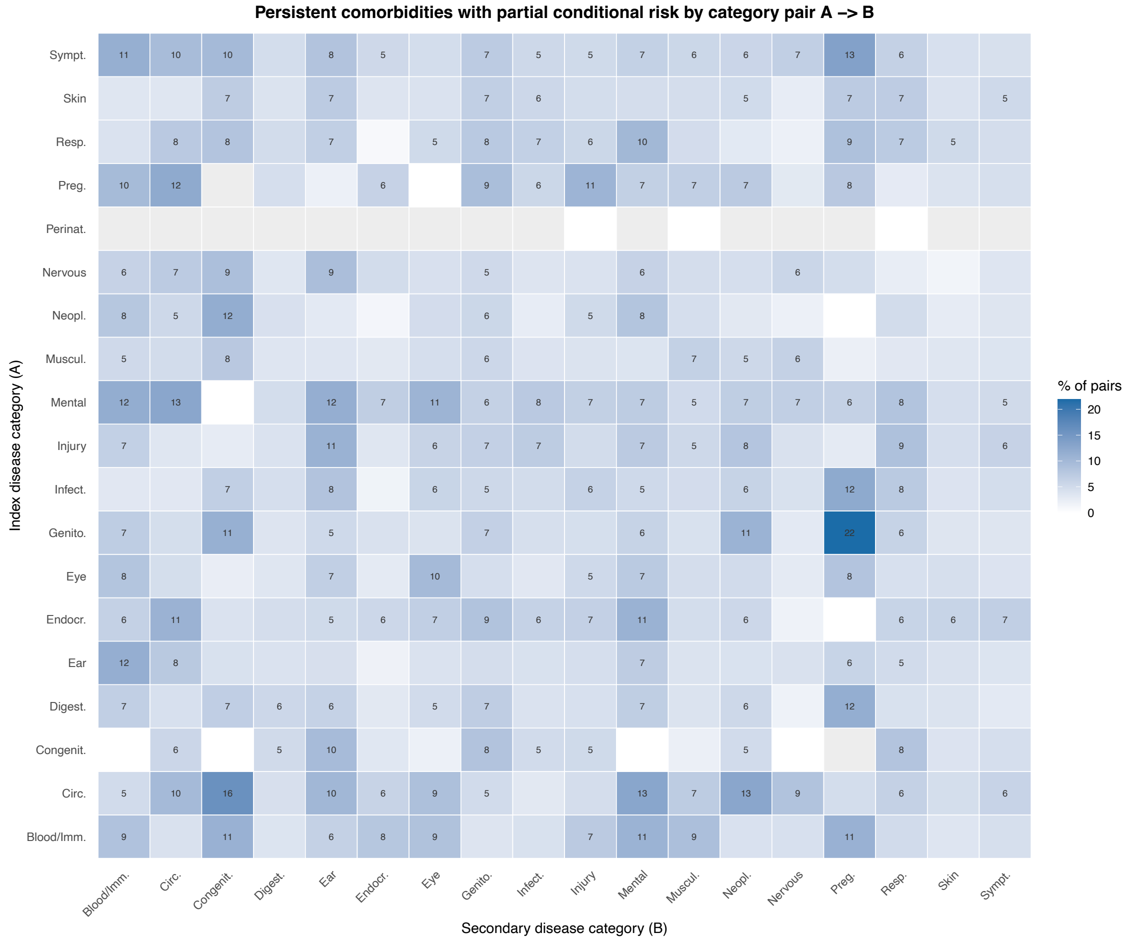


**Supplementary Figure 4.** Distribution of persistent cumulative comorbidity pairs with partial conditional risk across ICD-10 category pairs. These pairs are significant in all five cumulative windows and in at least one but not all four conditional windows. Genitourinary → Pregnancy (22%) and Mental → Circulatory (13%) show the highest proportions, consistent with sustained biological mechanisms linking these categories that generate detectable conditional risk across multiple time intervals.

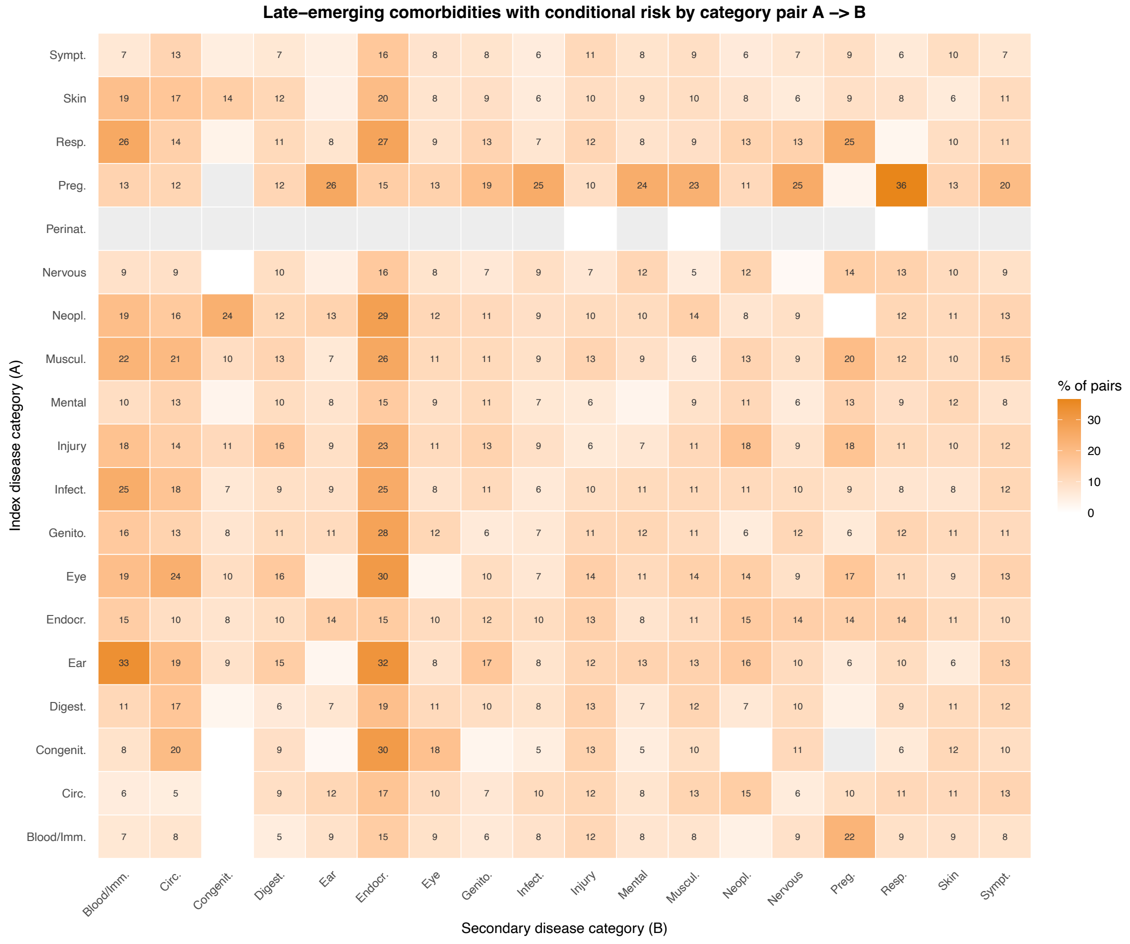


**Supplementary Figure 5.** Distribution of transient-persistent comorbidity pairs with conditional risk (significant only in longer cumulative windows, with additional conditional signal) across ICD-10 category pairs. Pregnancy → Respiratory (36%) shows the highest proportion, consistent with the long-term cardiometabolic and respiratory sequelae of obstetric conditions that emerge years after the index diagnosis. Ear → Blood/Immune (33%) and Ear → Endocrine/Metabolic (32%) also show elevated proportions, reflecting the progressive systemic associations of chronic ear conditions.

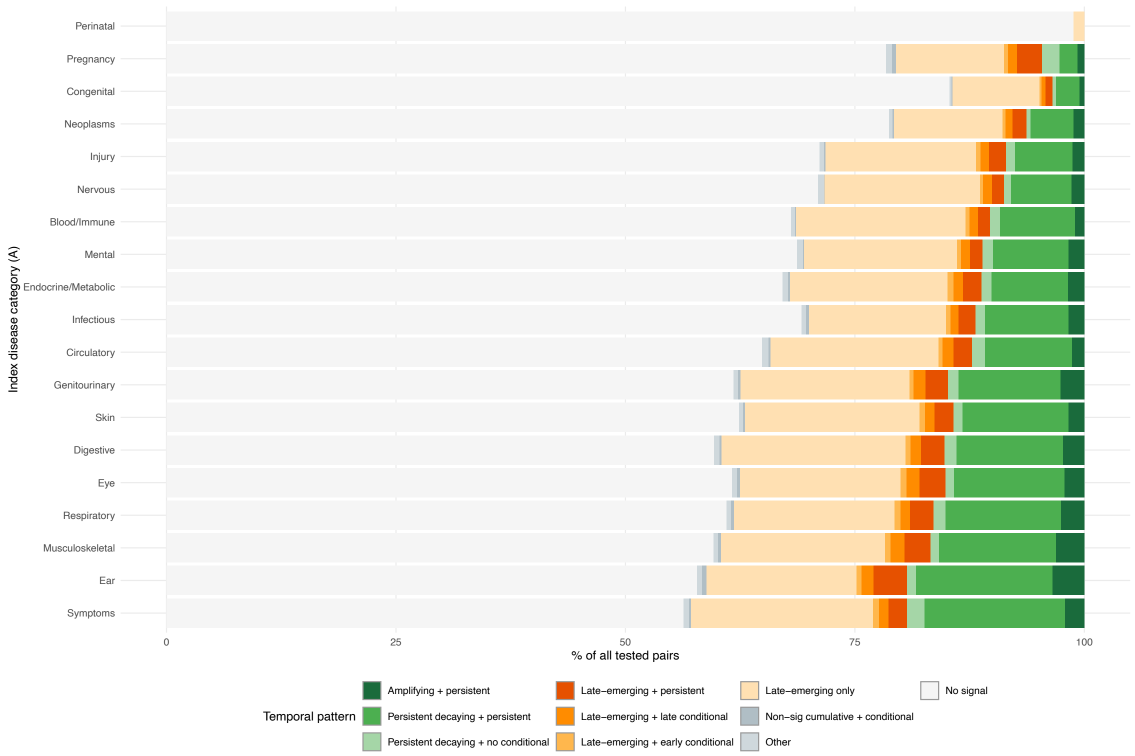


**Supplementary Figure 6. Full temporal taxonomy of comorbidity pairs by ICD-10 disease category.** Stacked bar chart showing the distribution of combined temporal patterns among all 419,995 pairs with data in all nine follow-up windows (five cumulative and four conditional), restricted to the index disease category. Green shades = pairs with robust cumulative signal (significant in all five cumulative windows): dark green = amplifying signal (positive cumulative slope, n = 8,604); medium green = persistent-decaying with persistent conditional risk (n = 42,457); light green = persistent-decaying without conditional risk (n = 4,780). Orange shades = pairs with late-emerging cumulative signal (significant in longer but not shorter cumulative windows): dark orange = late-emerging with persistent conditional risk (n = 8,978); medium orange = late-emerging with late conditional risk only (n = 4,557); light orange = late-emerging with early conditional risk only (n = 2,109); pale orange = late-emerging with no conditional signal (n = 72,468). Grey = non-significant cumulatively but with a conditional signal (n = 928). White = no significant signal in any window (n = 273,440, excluded from display to focus on pairs with detectable signal in at least one window). Significance threshold throughout: lfsr < 0.05, lower bound of the 95% credible interval ≥ 1.01, ≥100 cases. Analysis performed for the combined population (both sexes).

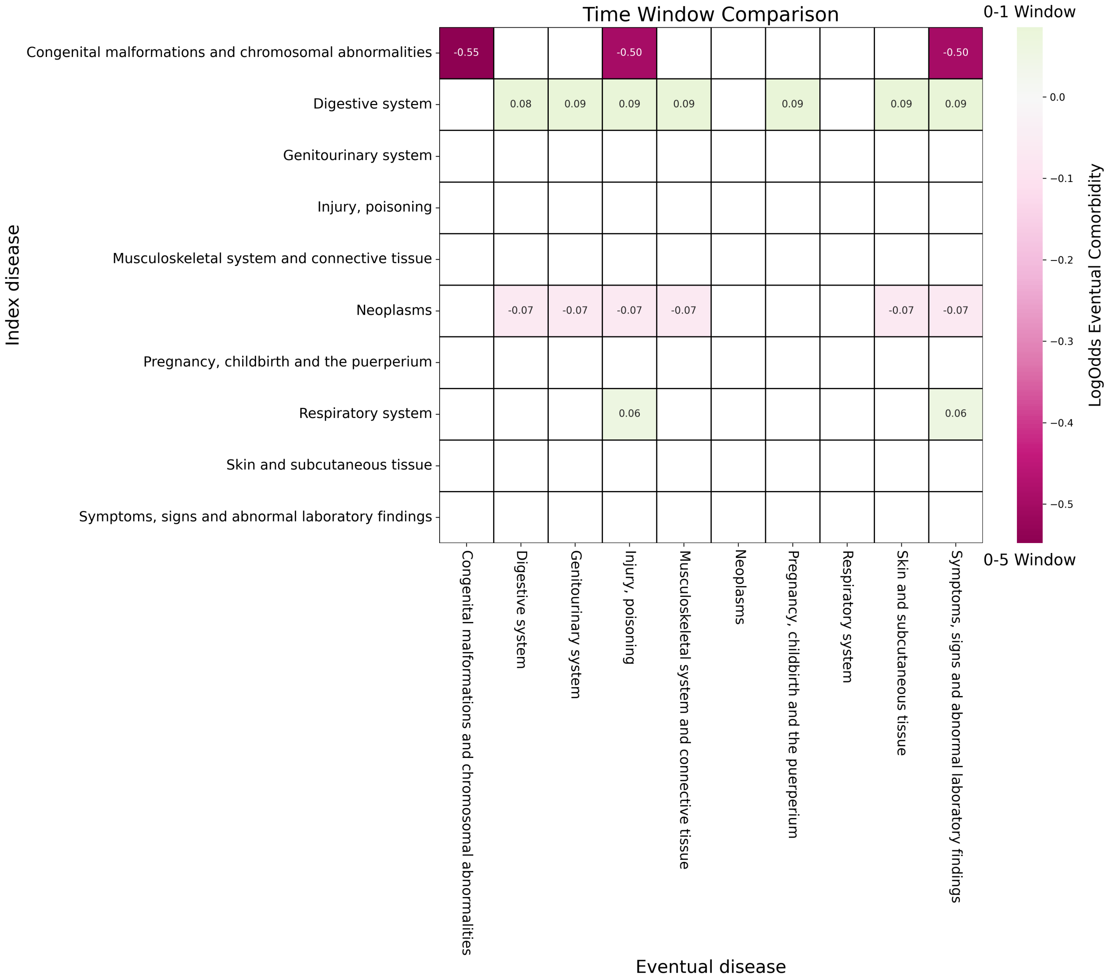


**Supplementary Figure 7. Eventual comorbidities comparison across time-windows.** Odds ratio of having a pathway from the index disease group (y-axis) to the corresponding eventual disease group (x-axis). Increased odds within the 1-year (5-year) window are coloured green (magenta).

**Supplementary Table 1.** Temporal trajectories of relative risk for the four paradigmatic examples of the biological clock patterns.

| | Episodic | Chronic stable | Chronic progressive | Transient-persistent |
|---|---|---|---|---|
| **Window** | **A63→A64** | **F10→K70** | **J01→J32** | **R06→J96** |
| Cumulative 0-1 | 24.30 (16.02–36.85) [n=135]* | 28.15 (22.49-35.23) [n=511]* | 29.80 (25.76-34.46) [n=1268]* | 12.53 (9.61-16.34) [n=195]* |
| Cumulative 0-2 | 18.93 (13.46–26.62) [n=161]* | 28.26 (23.50-33.98) [n=755]* | 20.54 (18.58-22.71) [n=1941]* | 9.08 (7.38-11.17) [n=267]* |
| Cumulative 0-3 | 15.45 (11.50–20.75) [n=184]* | 27.88 (23.74-32.74) [n=979]* | 16.90 (15.58-18.33) [n=2523]* | 7.50 (6.33-8.88) [n=354]* |
| Cumulative 0-4 | 11.50 (8.95–14.77) [n=209]* | 27.34 (23.67-31.58) [n=1195]* | 14.78 (13.78-15.86) [n=3050]* | 6.53 (5.66-7.54) [n=442]* |
| Cumulative 0-5 | 9.94 (7.92–12.48) [n=231]* | 25.54 (22.44-29.07) [n=1397]* | 12.50 (11.76-13.29) [n=3529]* | 5.87 (5.19-6.65) [n=553]* |
| Conditional 1-2 | 5.48 (2.66–11.31) [n=26] | 27.46 (19.90-37.89) [n=244]* | 12.99 (11.25-14.98) [n=673]* | 4.57 (3.30-6.35) [n=72] |
| Conditional 2-3 | 4.17 (2.38–7.32) [n=23] | 26.55 (19.11-36.89) [n=224]* | 10.60 (9.14-12.28) [n=582]* | 4.46 (3.35-5.95) [n=87] |
| Conditional 3-4 | 2.82 (1.78–4.47) [n=25] | 24.92 (17.99-34.52) [n=216]* | 9.39 (8.12-10.85) [n=527]* | 3.82 (2.89-5.03) [n=88] |
| Conditional 4-5 | 2.83 (1.65–4.86) [n=22] | 18.25 (13.57-24.55) [n=202]* | 6.35 (5.53-7.28) [n=479]* | 3.85 (2.98-4.96) [n=111]* |

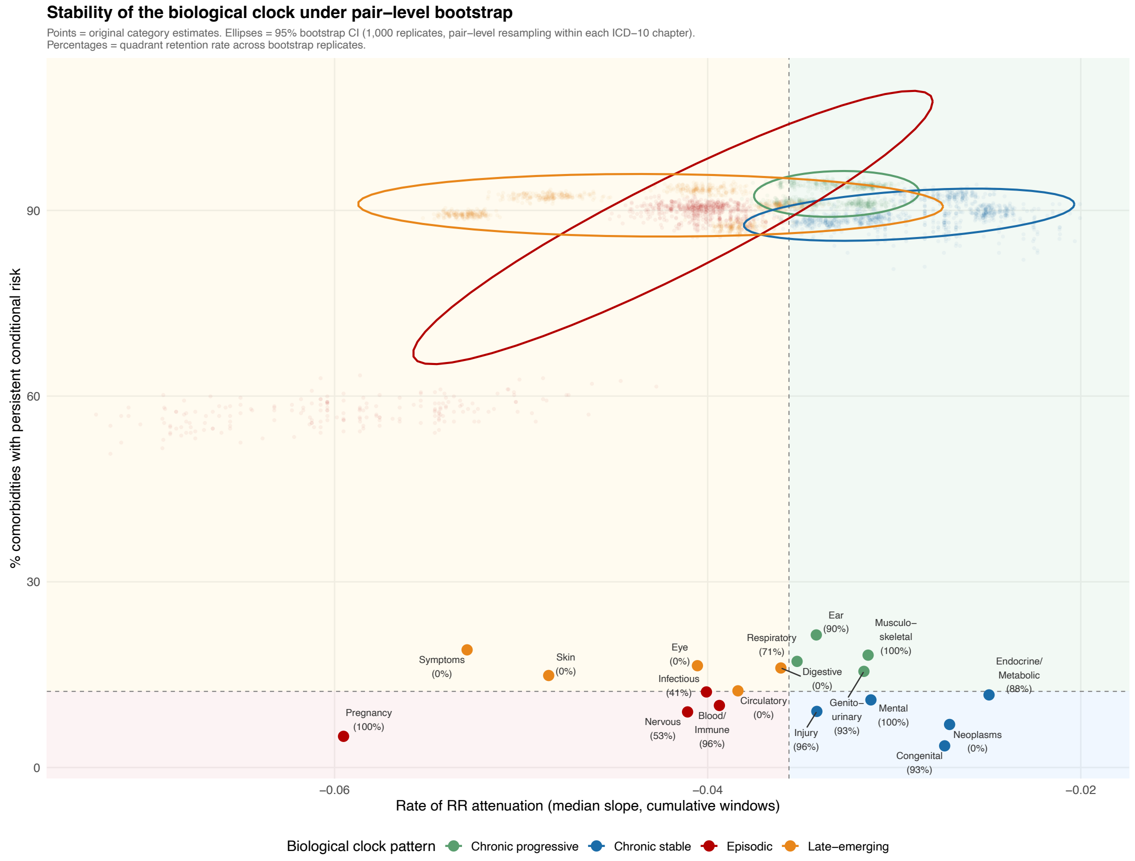


**Supplementary Figure 8. Stability of the biological clock under pair-level bootstrap resampling**. Each point represents the original category estimate in the two-dimensional clock space (rate of cumulative RR attenuation, x-axis; proportion of pairs with persistent conditional risk, y-axis). Ellipses show 95% bootstrap confidence intervals computed from 1,000 replicates in which disease pairs were resampled with replacement within each ICD-10 chapter and quadrant boundaries were recalculated as the medians of both dimensions in each replicate. Point labels show the category name and bootstrap quadrant retention rate (percentage of replicates retaining the original quadrant assignment). Background colours indicate quadrant: red = episodic; orange = transient-persistent; green = chronic progressive; blue = chronic stable. The faint point cloud shows 200 individual bootstrap replicates for each category.

**Supplementary Table 2. Robustness of the biological clock to pair-level resampling and leave-one-index-disease-out sensitivity analysis.** For each ICD-10 category, the table reports: the original quadrant assignment; the 95% bootstrap confidence interval for the median cumulative slope (slope CI) and for the proportion of pairs with persistent conditional risk (persistent risk CI), computed from 1,000 pair-level bootstrap replicates with recalculated quadrant boundaries; the bootstrap quadrant retention rate (% of replicates retaining the original quadrant); the leave-one-index-disease-out quadrant retention rate (% of single-disease-exclusion analyses retaining the original quadrant); the most influential index disease (the disease whose removal caused the largest shift in median slope); and the quadrant assigned by k-means clustering (k=4) on normalised clock coordinates. Categories are ordered by decreasing bootstrap retention rate.

| Category | Original quadrant | Slope CI | Persist CI | Boot retention | LOO retention | Most influential | Quadrant (kmeans) |
|---|---|---|---|---|---|---|---|
| **Musculoskeletal** | Chronic progressive | (-0.0329, -0.0300) | (93.3, 94.5) | 100 | 100 | M51 | Chronic progressive |
| **Pregnancy** | Episodic | (-0.0720, -0.0485) | (52.7, 61.5) | 100 | 0 | O04 | Episodic |
| **Mental** | Chronic stable | (-0.0331, -0.0297) | (87.4, 90.0) | 99.6 | 0 | F17 | Chronic stable |
| **Injury** | Chronic stable | (-0.0361, -0.0323) | (87.0, 89.3) | 96.1 | 0 | S83 | Chronic stable |
| **Blood/Immune** | Episodic | (-0.0430, -0.0366) | (86.7, 90.7) | 95.7 | 0 | D50 | Chronic stable |
| **Genitourinary** | Chronic progressive | (-0.0330, -0.0302) | (90.2, 91.8) | 92.7 | 100 | N30 | Chronic progressive |
| **Congenital** | Chronic stable | (-0.0343, -0.0211) | (83.5, 91.4) | 92.6 | 0 | Q40 | Chronic stable |
| **Ear** | Chronic progressive | (-0.0366, -0.0319) | (93.3, 95.3) | 90.2 | 92.3 | H61 | Chronic progressive |
| **Endocrine/Metabolic** | Chronic stable | (-0.0265, -0.0228) | (88.3, 91.0) | 87.6 | 0 | E78 | Chronic stable |
| **Respiratory** | Chronic progressive | (-0.0372, -0.0336) | (90.4, 92.2) | 71 | 78.4 | J03 | Chronic progressive |
| **Nervous** | Episodic | (-0.0442, -0.0381) | (88.8, 92.0) | 53 | 0 | G56 | Chronic stable |
| **Infectious** | Episodic | (-0.0421, -0.0382) | (89.6, 91.5) | 41.3 | 0 | A09 | Chronic stable |
| **Neoplasms** | Chronic stable | (-0.0301, -0.0249) | (91.0, 93.7) | 0.5 | 0 | D22 | Chronic stable |
| **Circulatory** | Transient-persistent | (-0.0397, -0.0367) | (86.3, 88.6) | 0 | 0 | I10 | Chronic stable |
| **Digestive** | Transient-persistent | (-0.0373, -0.0344) | (90.0, 91.5) | 0 | 0 | K59 | Chronic progressive |
| **Eye** | Transient-persistent | (-0.0427, -0.0379) | (92.4, 94.4) | 0 | 0 | H52 | Chronic progressive |
| **Skin** | Transient-persistent | (-0.0510, -0.0463) | (91.5, 93.0) | 0 | 0 | L70 | Transient-persistent |
| **Symptoms** | Transient-persistent | (-0.0544, -0.0514) | (88.7, 90.0) | 0 | 0 | R73 | Transient-persistent |

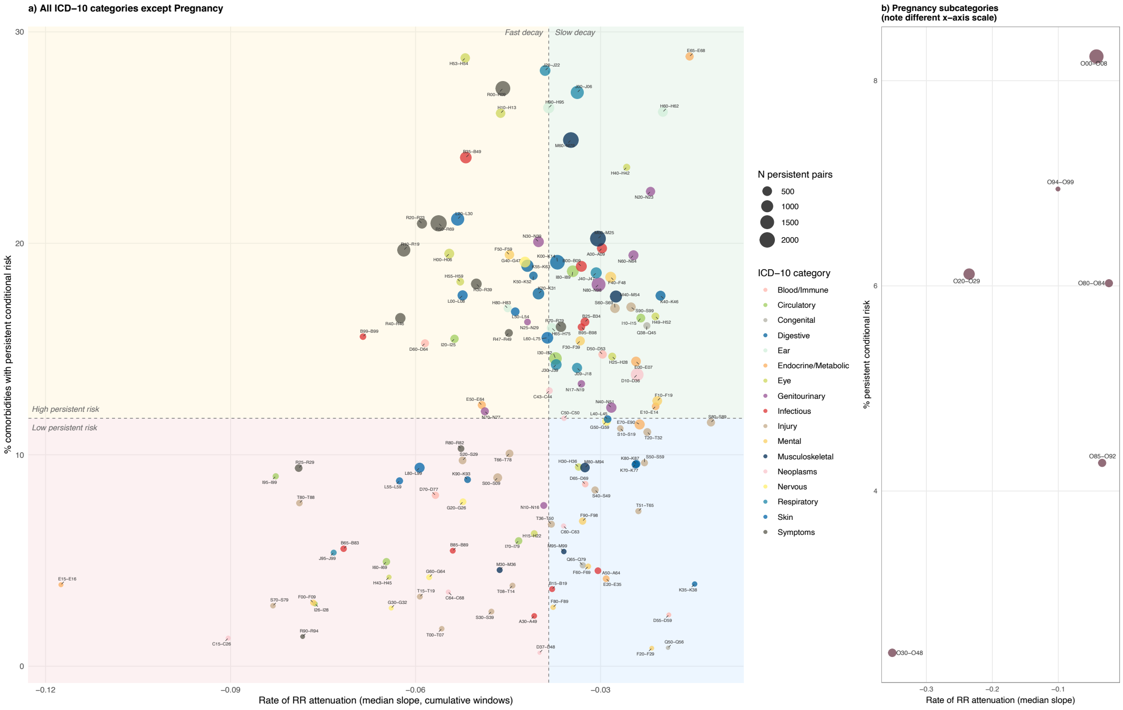


**Supplementary Figure 9. Two-dimensional biological clock at ICD-10 subcategory level.** Each point represents one ICD-10 subcategory restricted to those with at least 10 persistent comorbidity pairs (significant in all five cumulative windows; n = 141 subcategories across 18 chapters). The x-axis shows the median weighted slope of the posterior relative risk across the five cumulative follow-up windows (0-1 to 0-5 years); the y-axis shows the proportion of comorbidity pairs in that subcategory with persistent conditional risk (significant associations in at least one early [years 1-2 or 2-3] and at least one late [years 3-4 or 4-5] conditional window). Quadrant boundaries are defined by the medians of both dimensions across all subcategories. Point size is proportional to the number of persistent comorbidity pairs. Colour indicates ICD-10 chapter. Panel (a) shows all categories except Pregnancy; panel (b) shows Pregnancy subcategories on a separate x-axis scale (slope range -0.35 to -0.02) due to their markedly faster cumulative attenuation relative to other categories (slope range -0.12 to -0.01). Background colours indicate quadrant: red = episodic (fast decay, low persistent risk); orange = transient-persistent (fast decay, high persistent risk); green = chronic progressive (slow decay, high persistent risk); blue = chronic stable (slow decay, low persistent risk). Significance threshold throughout: lfsr < 0.05, lower bound of the 95% credible interval ≥ 1.01, ≥100 cases.

**Supplementary Table 3. Within-category heterogeneity of biological clock dynamics at the ICD-10 subcategory level.** For each ICD-10 chapter, the table reports: the number of subcategories with sufficient data (≥10 persistent comorbidity pairs); the range, minimum, and maximum of the median cumulative attenuation slope across subcategories; the range, minimum, and maximum of the proportion of subcategories with persistent conditional risk; the number of distinct biological clock quadrants occupied by subcategories within that chapter; and the names of those quadrants. Categories are ordered by decreasing slope range, representing decreasing within-category heterogeneity in the cumulative attenuation dimension.

| Category | subcategories | Slope range | Slope (min) | Slope (max) | Persist range | Persist (min) | Persist (max) | quadrants | quadrants |
|---|---|---|---|---|---|---|---|---|---|
| Pregnancy | 6 | 0.3285 | -0.3518 | -0.0233 | 5.8 | 2.4 | 8.2 | 2 | Chronic stable, Episodic |
| Endocrine/Metabolic | 7 | 0.1019 | -0.1175 | -0.0156 | 25 | 3.9 | 28.8 | 4 | All |
| Injury | 18 | 0.071 | -0.0831 | -0.0121 | 15.2 | 1.8 | 17 | 3 | Chronic progressive, Chronic stable, Episodic |
| Neoplasms | 7 | 0.0663 | -0.0904 | -0.0241 | 13.2 | 0.6 | 13.8 | 3 | Chronic progressive, Chronic stable, Episodic |
| Circulatory | 8 | 0.0592 | -0.0827 | -0.0235 | 15.7 | 2.9 | 18.7 | 3 | Chronic progressive, Transient-persistent, Episodic |
| Mental | 9 | 0.0558 | -0.0766 | -0.0208 | 18.6 | 0.9 | 19.5 | 4 | All |
| Eye | 10 | 0.0432 | -0.0643 | -0.0211 | 24.6 | 4.2 | 28.8 | 4 | All |
| Respiratory | 6 | 0.0426 | -0.0733 | -0.0307 | 22.8 | 5.4 | 28.2 | 3 | Chronic progressive, Transient-persistent, Episodic |
| Symptoms | 11 | 0.0425 | -0.079 | -0.0364 | 25.9 | 1.4 | 27.3 | 3 | Chronic progressive, Transient-persistent, Episodic |
| Infectious | 11 | 0.0419 | -0.0717 | -0.0298 | 21.7 | 2.4 | 24.1 | 4 | All |
| Blood/Immune | 5 | 0.0395 | -0.0585 | -0.0189 | 12.8 | 2.4 | 15.3 | 4 | All |
| Digestive | 9 | 0.0368 | -0.0516 | -0.0147 | 15.2 | 3.9 | 19.1 | 4 | All |
| Nervous | 5 | 0.0349 | -0.064 | -0.029 | 16.4 | 2.8 | 19.1 | 3 | Chronic stable, Transient-persistent, Episodic |
| Skin | 7 | 0.0338 | -0.0626 | -0.0289 | 12.4 | 8.8 | 21.1 | 3 | Chronic stable, Transient-persistent, Episodic |
| Genitourinary | 9 | 0.0269 | -0.0488 | -0.0219 | 14.9 | 7.6 | 22.5 | 3 | Chronic progressive, Transient-persistent, Episodic |
| Ear | 4 | 0.0252 | -0.0451 | -0.0199 | 10.4 | 16 | 26.4 | 2 | Chronic progressive, Transient-persistent |
| Musculoskeletal | 6 | 0.0189 | -0.0464 | -0.0275 | 20.3 | 4.5 | 24.9 | 3 | Chronic progressive, Chronic stable, Episodic |
| Congenital | 3 | 0.0138 | -0.0329 | -0.0191 | 15.2 | 0.9 | 16.1 | 2 | Chronic progressive, Chronic stable |

**Supplementary Table 4. Category-level sex differences in age at first diagnosis and directional enrichment analysis.** For each ICD-10 disease category, the table reports the number of diseases included (number of diseases), the estimated mean age difference in years between women and men (beta (years); women minus men), 95% confidence intervals (CI (low) - CI(high)), and FDR-adjusted P values (FDR). Positive values indicate later diagnosis in women, and negative values indicate later diagnosis in men. The table additionally provides results from the sensitivity analysis restricted to diseases with significant sex differences, including the number of significant diseases per category diagnosed later in women (n (LiW) and men (n (LiM)), and corresponding FDR-adjusted P values from directional binomial tests (FDR (W), FDR (M)).

| **disease category** | **Number of diseases** | **beta (years)** | **CI (low)** | **CI (high)** | **FDR** | **n (LiW)** | **n (LiM)** | **FDR (W)** | **FDR (M)** |
|---|---|---|---|---|---|---|---|---|---|
| Injury, poisoning | 187 | 7.18 | 6.52 | 7.84 | 2.89e-86 | 112 | 8 | 1.22e-23 | 1 |
| Circulatory system | 77 | 4.2 | 3.62 | 4.79 | 1.72e-41 | 58 | 5 | 7.48e-12 | 1 |
| Musculoskeletal system and connective tissue | 79 | 2.44 | 2.03 | 2.85 | 4.62e-29 | 55 | 9 | 1.06e-08 | 1 |
| Respiratory system | 63 | 2.74 | 2.28 | 3.21 | 4.92e-29 | 33 | 6 | 1.28e-05 | 1 |
| Endocrine, nutritional and metabolic | 67 | 3.22 | 2.56 | 3.89 | 2.16e-20 | 21 | 15 | 0.227 | 1 |
| Symptoms, signs and abnormal laboratory findings | 88 | 2.05 | 1.61 | 2.48 | 5.51e-19 | 52 | 17 | 2.39e-05 | 1 |
| Blood and blood-forming organs (immune) | 33 | -6.5 | -8.12 | -4.88 | 1.73e-14 | 11 | 8 | 0.342 | 1 |
| Infectious and parasitic | 145 | 2.36 | 1.75 | 2.96 | 7.65e-14 | 49 | 16 | 3.81e-05 | 1 |
| Digestive system | 70 | 1.83 | 1.27 | 2.38 | 4.14e-10 | 46 | 11 | 4.25e-06 | 1 |
| Skin and subcutaneous tissue | 71 | 2.01 | 1.37 | 2.66 | 2.068e-09 | 43 | 6 | 1.28e-07 | 1 |
| Eye and adnexa | 47 | 2.01 | 1.33 | 2.69 | 1.36e-08 | 32 | 4 | 2.91e-06 | 1 |
| Genitourinary system | 73 | -2.12 | -2.91 | -1.32 | 3.16e-07 | 16 | 21 | 0.837 | 1 |
| Ear and mastoid process | 24 | 1.8 | 0.96 | 2.63 | 3.51e-05 | 14 | 1 | 0.00067 | 1 |
| Nervous system | 64 | 1.95 | 0.92 | 2.98 | 0.00028 | 29 | 4 | 1.09e-05 | 1 |
| Congenital malformations and chromosomal abnormalities | 82 | 4.62 | 1.66 | 7.59 | 0.00274 | 30 | 4 | 6.93e-06 | 1 |
| Mental and behavioural | 78 | 0.99 | 0.35 | 1.63 | 0.00274 | 47 | 8 | 1.45e-07 | 1 |
| Neoplasms | 125 | 0.79 | -0.18 | 1.75 | 0.116 | 35 | 23 | 0.088 | 1 |
| Certain conditions originating in the perinatal period | 44 | 5.33 | -9.03 | 19.7 | 0.466 | 12 | 2 | 0.0083 | 1 |

**Supplementary Table 5. Multimorbidity initiators and sinks identified by weighted outdegree–PageRank ranking discrepancy.** Diseases are classified as initiators (weighted outdegree ranking substantially exceeds PageRank ranking) or sinks (PageRank ranking substantially exceeds weighted outdegree ranking) based on a z-score threshold of |z| > 2 consistent across at least two of three weighting schemes (posterior shrunk relative risk, number of incident secondary cases, and their product). z-mean: mean z-score across the three weighting schemes. Consistency: number of weighting schemes in which the disease exceeded the |z| > 2 threshold. Role: I for initiation and S for sink. Diseases marked with * are likely coding artefacts and are excluded from the main text interpretation. Analysis performed on the combined population (both sexes) 0-5-year comorbidity network (n = 845 diseases after exclusion of three high-prevalence diseases: M54, J00, T14).

| ICD10 | Disease | Role | z-mean | Consistency |
|---|---|---|---|---|
| I10 | Essential hypertension | I | 3.99 | 3/3 |
| C61 | Malignant neoplasm of prostate | I | 3.48 | 3/3 |
| D56 | Thalassaemia | I | 3.42 | 3/3 |
| B18 | Chronic viral hepatitis | I | 2.99 | 2/3 |
| M81 | Osteoporosis without pathological fracture | I | 2.79 | 3/3 |
| C18 | Malignant neoplasm of colon | I | 2.71 | 3/3 |
| I21 | Acute myocardial infarction | I | 2.55 | 3/3 |
| B19 | Unspecified viral hepatitis | I | 2.46 | 3/3 |
| T65 | Toxic effects of other substances | I | 2.32 | 2/3 |
| I61 | Intracerebral haemorrhage | I | 2.24 | 2/3 |
| E05 | Thyrotoxicosis | I | 2.22 | 2/3 |
| K26 | Duodenal ulcer | I | 2.19 | 2/3 |
| C50 | Malignant neoplasm of breast | I | 2.11 | 2/3 |
| B16 | Acute hepatitis B | I | 2.05 | 2/3 |
| R71 | Abnormal red blood cells | I | 2 | 2/3 |
| T11 | Injuries of other parts of body (level unspecified) | I | 1.99 | 2/3 |
| T01 | Open wounds involving multiple body regions | I | 1.91 | 2/3 |
| H50 | Other strabismus | I | 1.8 | 2/3 |
| F30 | Maniac episodes | I | 1.23 | 2/3 |
| K72 | Hepatic failure | S | -6.61 | 3/3 |
| C79 | Secondary malignant neoplasm (other sites) | S | -4.63 | 3/3 |
| C78 | Secondary malignant neoplasm (respiratory/digestive organs) | S | -4.28 | 3/3 |
| B78 | Strongyloidiasis* | S | -4.13 | 3/3 |
| T00 | Superficial injuries involving multiple body regions* | S | -4.07 | 3/3 |
| N32 | Other disorders of bladder | S | -3.41 | 3/3 |
| C22 | Malignant neoplasm of liver and intrahepatic bile ducts | S | -3.36 | 3/3 |
| E26 | Hyperaldosteronism | S | -3.26 | 2/3 |
| L97 | Ulcer of lower limb (non-decubitus) | S | -3.18 | 3/3 |
| L89 | Pressure ulcer | S | -3.18 | 3/3 |

| | | | | |
|---|---|---|---|---|
| D45 | Polycythaemia vera | S | -2.68 | 3/3 |
| J46 | Status asthmaticus | S | -2.66 | 3/3 |
| E20 | Hypoparathyroidism | S | -2.65 | 2/3 |
| R99 | Ill-defined and unknown cause of mortality | S | -2.56 | 2/3 |
| C81 | Hodgkin lymphoma | S | -2.46 | 2/3 |
| G62 | Other polyneuropathies | S | -2.38 | 3/3 |
| N50 | Other disorders of male genital organs | S | -2.34 | 2/3 |
| E55 | Vitamin D deficiency | S | -2.2 | 2/3 |
| J69 | Pneumonitis due to solids and liquids | S | -2.1 | 2/3 |
| I77 | Other disorders of arteries and arterioles | S | -1.99 | 2/3 |
| L28 | Lichen simplex chronicus and prurigo | S | -1.95 | 2/3 |
| G93 | Other disorders of brain | S | -1.72 | 2/3 |

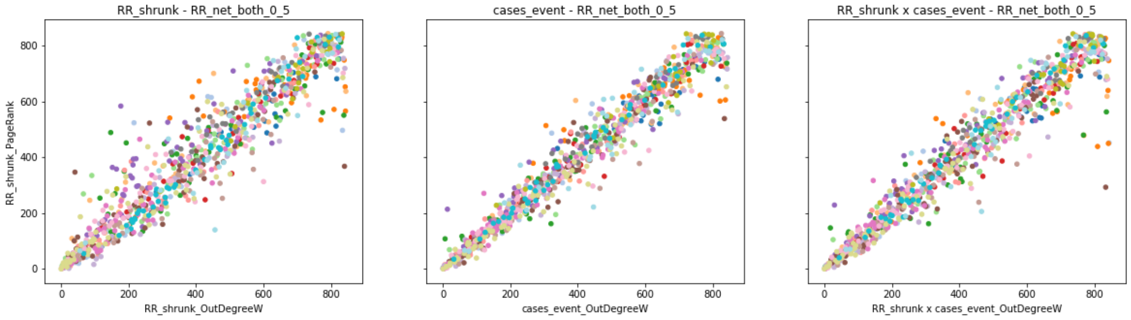


**Supplementary Figure 10. Network centrality discrepancy identifies multimorbidity initiators and sinks. Each point represents one ICD-10 disease.** The x-axis shows the weighted outdegree ranking (position 0 = highest weighted outdegree) and the y-axis shows the PageRank ranking (position 0 = highest PageRank), both computed on the 0–5-year combined-population comorbidity network. Three weighting schemes are shown: RR_shrunk (posterior relative risk), cases_event (number of incident cases), and RR_shrunk × cases_event (combined weight). Points above the diagonal represent diseases whose weighted outdegree ranking exceeds their PageRank ranking (multimorbidity initiators – high direct comorbidity burden with limited network propagation); points below the diagonal represent diseases whose PageRank ranking exceeds their weighted outdegree ranking (multimorbidity sinks – convergent destinations of multiple upstream trajectories). Colours indicate ICD-10 disease category. The red line shows the linear regression fit. Diseases with |z| > 2 that are consistent across at least two of the three weighting schemes are annotated (Supplementary Table 5).

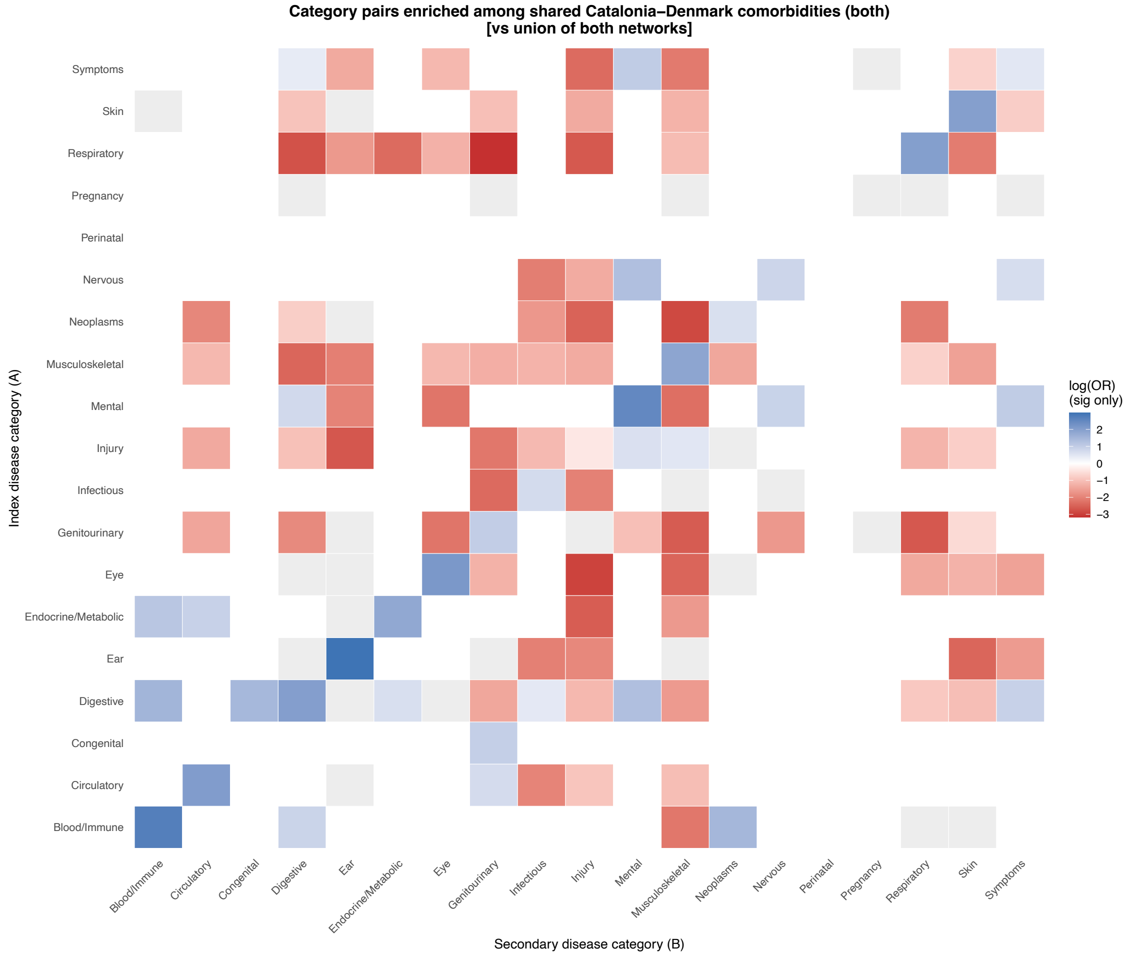


**Supplementary Figure 11.** Enrichment of ICD-10 category pairs among shared Catalonia–Denmark comorbidities (combined population, window 0–5 years). Each cell shows the log odds ratio from a Fisher's exact test comparing the proportion of pairs in that category combination among shared pairs versus Catalonia-only pairs. Blue = overrepresented among shared pairs; red = underrepresented; white = not significant after BH correction. Only significant associations (BH-adjusted p ≤ 0.05) are coloured.

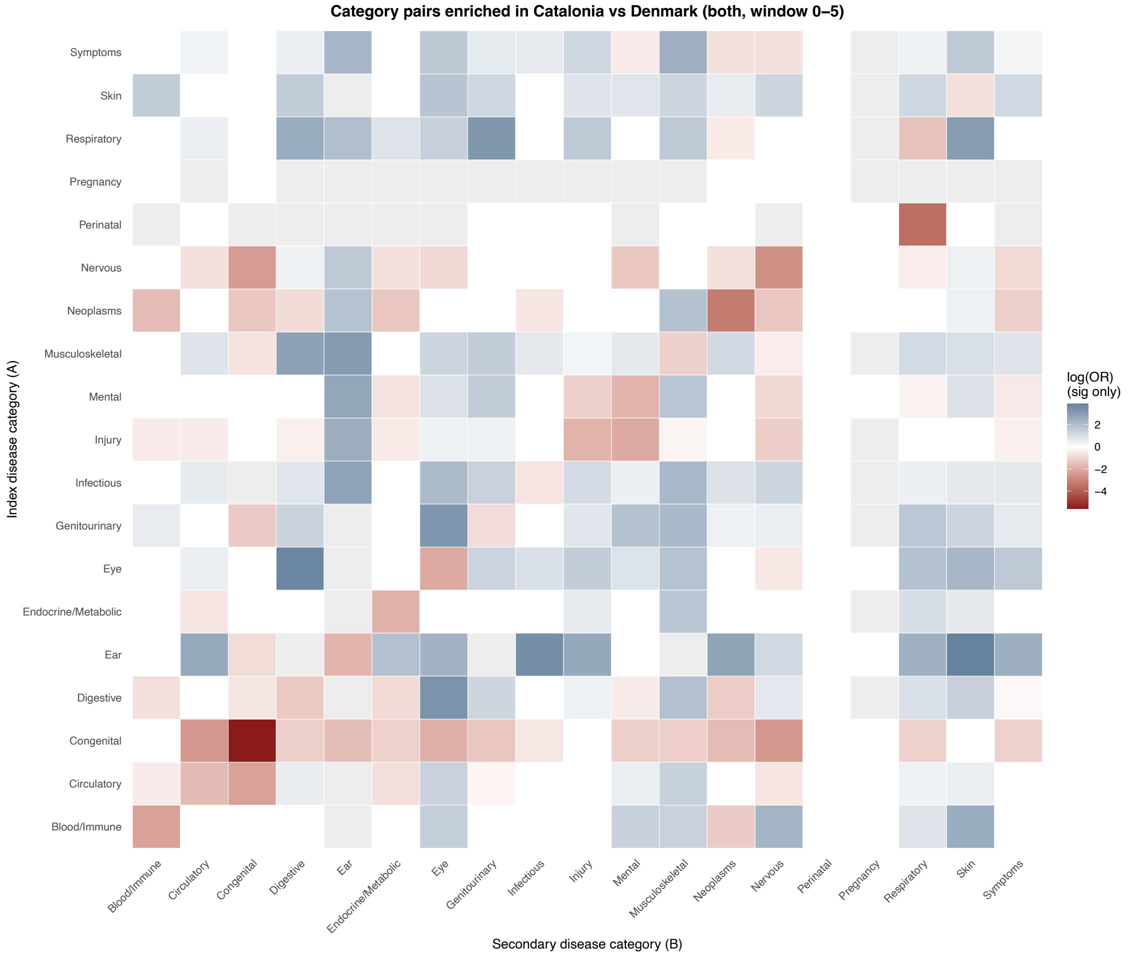


**Supplementary Figure 12.** Differential representation of ICD-10 category pairs in Catalonia versus Denmark. Each cell shows the log-odds ratio comparing the proportion of pairs in that category combination among all significant associations in Catalan versus all significant associations in Danish, restricted to the common disease universe. Blue = overrepresented in Catalonia; red = overrepresented in Denmark; white = not significant. Results shown for the combined population.

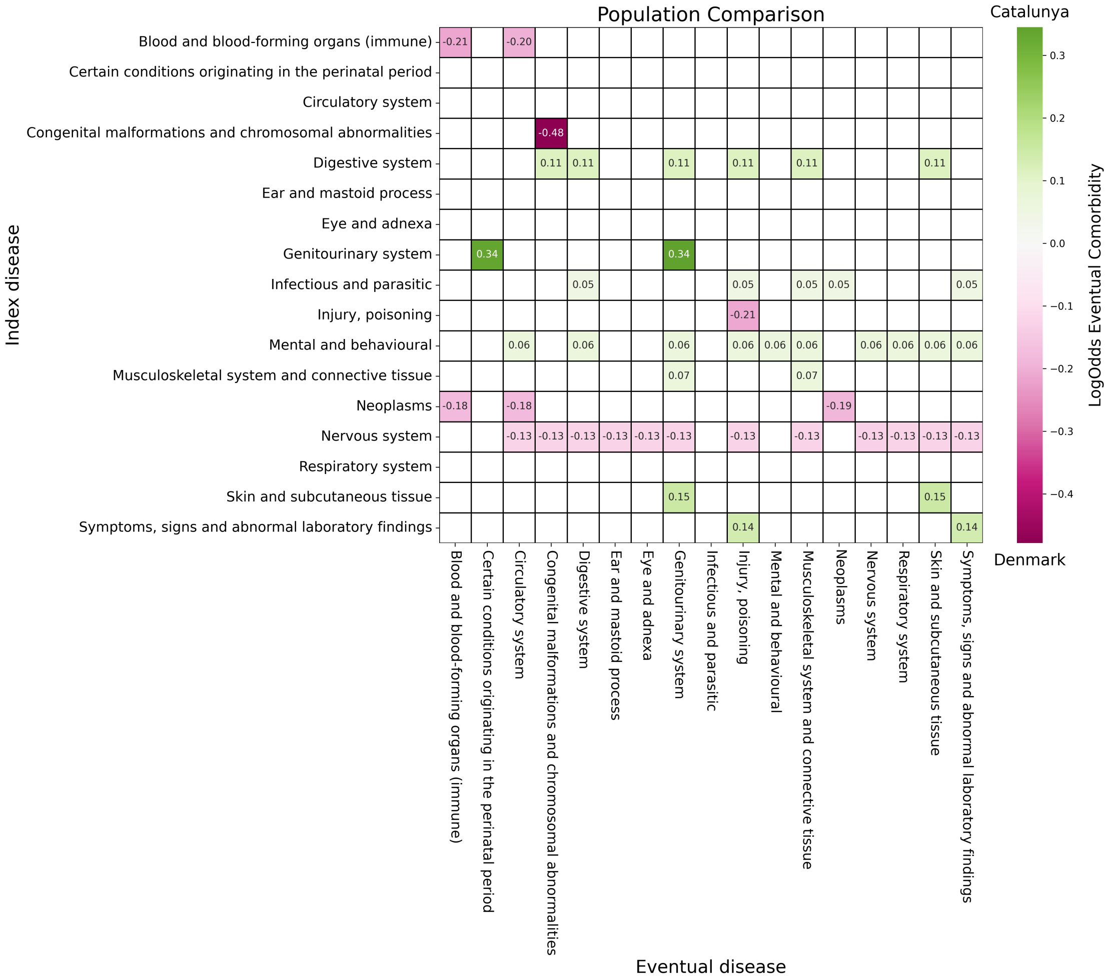


**Supplementary Figure 13. Eventual comorbidities comparison across health systems.** Odds ratio of having a pathway from the index disease group (y-axis) to the corresponding eventual disease group (x-axis). Increased odds within the Catalan (Danish) population are colored green (magenta).

**Supplementary Table 6.** Diseases with significant differences in prevalence between women and men in Catalonia and Denmark (significantly more prevalent in women in one population and in men in the other, and vice versa).

| Disease | D (W) | D (M) | D (OR) | C (W) | C (M) | C (OR) |
|---|---|---|---|---|---|---|
| Hookworm diseases | 458 | 360 | 1.367 | 87 | 165 | 0.504 |
| Malignant neoplasm of palate | 541 | 436 | 1.333 | 92 | 128 | 0.687 |
| Malignant neoplasm of anus and anal canal | 2102 | 1035 | 2.183 | 455 | 607 | 0.716 |
| Malignant neoplasm of other and ill-defined sites | 4665 | 3468 | 1.446 | 169 | 214 | 0.755 |
| Secondary and unspecified malignant neoplasm of lymph nodes | 22049 | 17295 | 1.372 | 797 | 994 | 0.766 |
| Carcinoma in situ of other and unspecified genital organs | 4966 | 1270 | 4.206 | 356 | 926 | 0.367 |
| Other disorders of psychological development | 185 | 105 | 1.893 | 219 | 326 | 0.642 |
| Chronic hepatitis, not elsewhere classified | 2987 | 1752 | 1.832 | 1611 | 2333 | 0.66 |
| Osteopathies in diseases classified elsewhere | 1408 | 1086 | 1.393 | 280 | 358 | 0.747 |
| Other abnormal immunological findings in serum | 1721 | 1294 | 1.429 | 3061 | 15829 | 0.184 |
| Glycosuria | 576 | 159 | 3.893 | 250 | 401 | 0.596 |
| Chlamydia psittaci infection | 116 | 199 | 0.626 | 76 | 32 | 2.27 |
| Other rickettsioses | 97 | 175 | 0.595 | 162 | 109 | 1.421 |
| Respiratory conditions due to inhalation of chemicals, gases, fumes and vapours | 952 | 1490 | 0.686 | 1621 | 1131 | 1.37 |
| Neuromuscular dysfunction of bladder, not elsewhere classified | 8993 | 13817 | 0.698 | 1556 | 992 | 1.499 |
| Transitory neonatal disorders of calcium and magnesium metabolism | 373 | 524 | 0.764 | 26 | 12 | 2.071 |
| Other conditions of integument specific to fetus and newborn | 686 | 5967 | 0.123 | 108 | 57 | 1.811 |
| Crushing injury and traumatic amputation of part of abdomen, lower back and pelvis | 170 | 265 | 0.689 | 821 | 73 | 10.752 |
| Injury of blood vessels at lower leg level | 608 | 852 | 0.766 | 27 | 10 | 2.581 |
| Burn and corrosion of wrist and hand | 20284 | 30075 | 0.723 | 3692 | 2319 | 1.522 |
| Asphyxiation | 873 | 1235 | 0.759 | 508 | 313 | 1.551 |
| Effects of other external causes | 7289 | 17666 | 0.442 | 1171 | 851 | 1.315 |
| Complications of cardiac and vascular prosthetic devices, implants and grafts | 2928 | 4743 | 0.663 | 201 | 124 | 1.549 |
| Complications of genitourinary prosthetic devices, implants and grafts | 7082 | 24883 | 0.304 | 292 | 161 | 1.734 |

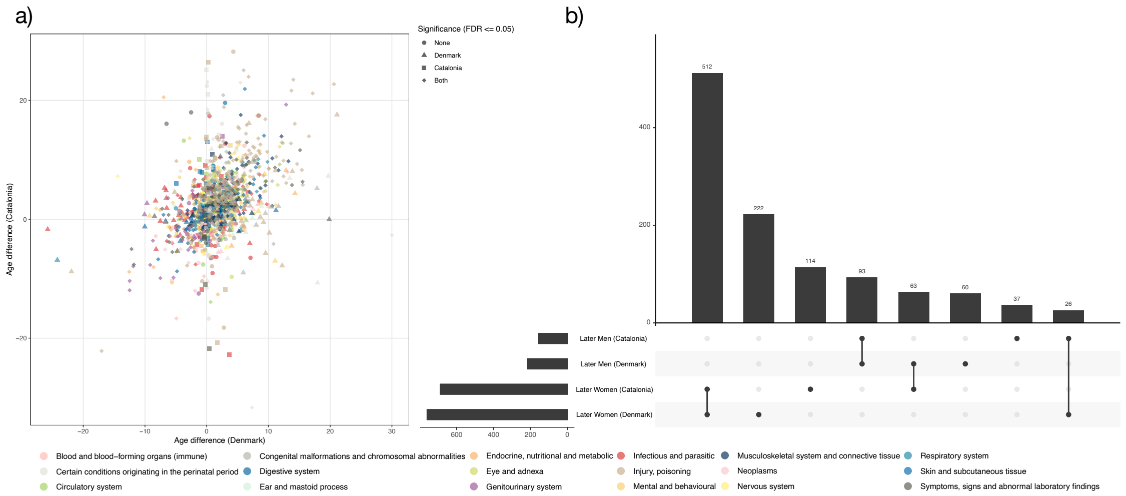


**Supplementary Figure 14. Cross-cohort comparison of sex differences in age at first diagnosis between Catalonia and Denmark**. A) Scatterplot of disease-specific mean age differences (women – men) in Denmark (x-axis) versus Catalonia (y-axis). Each point represents one ICD-10 three-digit disease and is coloured by ICD-10 category. Positive values indicate later diagnosis in women. Point shape indicates statistical significance (FDR ≤ 0.05) in Denmark, Catalonia, both, or neither. Clustering along the diagonal reflects cross-cohort concordance in effect direction. B) UpSet plot quantifying overlap of diseases with significant sex differences in age at diagnosis across populations. Bars indicate the number of diseases significant only in Denmark (n = 222), only in Catalonia (n = 93), in both (n = 114), or in neither cohort (n = 512).

**Supplementary Table 2.** Summary of the four biological clock patterns identified in the Catalan primary care comorbidity network. PC stands for Persistent Conditions, LE for Late emerging, and ER for Early risk.

| Pattern | ICD-10 categories | N (PC) | N (LE) | N (ER) | Clinical example |
|---|---|---|---|---|---|
| Episodic | Blood/Immune, Infectious, Nervous, Pregnancy | 6,243 | 701 | 581 | A63 → A64 |
| Chronic stable | Congenital, Endocrine/Metabolic, Injury, Mental, Neoplasms | 10,363 | 1,185 | 1,057 | F10 → K70 |
| Chronic progressive | Ear, Musculoskeletal, Genitourinary, Respiratory | 18,223 | 1,637 | 1,268 | J01 → J32 |
| Transient-persistent | Circulatory, Digestive, Eye, Skin, Symptoms | 24,815 | 2,123 | 1,979 | R06 → J96 |

**Supplementary Table 6.** Diseases with significant differences in prevalence between women and men in Catalonia and Denmark (significantly more prevalent in women in one population and in men in the other, and vice versa).

| Disease | D (W) | D (M) | D (OR) | C (W) | C (M) | C (OR) |
|---|---|---|---|---|---|---|
| Hookworm diseases | 458 | 360 | 1.367 | 87 | 165 | 0.504 |
| Malignant neoplasm of palate | 541 | 436 | 1.333 | 92 | 128 | 0.687 |
| Malignant neoplasm of anus and anal canal | 2102 | 1035 | 2.183 | 455 | 607 | 0.716 |
| Malignant neoplasm of other and ill-defined sites | 4665 | 3468 | 1.446 | 169 | 214 | 0.755 |
| Secondary and unspecified malignant neoplasm of lymph nodes | 22049 | 17295 | 1.372 | 797 | 994 | 0.766 |
| Carcinoma in situ of other and unspecified genital organs | 4966 | 1270 | 4.206 | 356 | 926 | 0.367 |
| Other disorders of psychological development | 185 | 105 | 1.893 | 219 | 326 | 0.642 |
| Chronic hepatitis, not elsewhere classified | 2987 | 1752 | 1.832 | 1611 | 2333 | 0.66 |
| Osteopathies in diseases classified elsewhere | 1408 | 1086 | 1.393 | 280 | 358 | 0.747 |
| Other abnormal immunological findings in serum | 1721 | 1294 | 1.429 | 3061 | 15829 | 0.184 |
| Glycosuria | 576 | 159 | 3.893 | 250 | 401 | 0.596 |
| Chlamydia psittaci infection | 116 | 199 | 0.626 | 76 | 32 | 2.27 |
| Other rickettsioses | 97 | 175 | 0.595 | 162 | 109 | 1.421 |
| Respiratory conditions due to inhalation of chemicals, gases, fumes and vapours | 952 | 1490 | 0.686 | 1621 | 1131 | 1.37 |
| Neuromuscular dysfunction of bladder, not elsewhere classified | 8993 | 13817 | 0.698 | 1556 | 992 | 1.499 |
| Transitory neonatal disorders of calcium and magnesium metabolism | 373 | 524 | 0.764 | 26 | 12 | 2.071 |
| Other conditions of integument specific to fetus and newborn | 686 | 5967 | 0.123 | 108 | 57 | 1.811 |
| Crushing injury and traumatic amputation of part of abdomen, lower back and pelvis | 170 | 265 | 0.689 | 821 | 73 | 10.752 |
| Injury of blood vessels at lower leg level | 608 | 852 | 0.766 | 27 | 10 | 2.581 |
| Burn and corrosion of wrist and hand | 20284 | 30075 | 0.723 | 3692 | 2319 | 1.522 |
| Asphyxiation | 873 | 1235 | 0.759 | 508 | 313 | 1.551 |
| Effects of other external causes | 7289 | 17666 | 0.442 | 1171 | 851 | 1.315 |
| Complications of cardiac and vascular prosthetic devices, implants and grafts | 2928 | 4743 | 0.663 | 201 | 124 | 1.549 |
| Complications of genitourinary prosthetic devices, implants and grafts | 7082 | 24883 | 0.304 | 292 | 161 | 1.734 |
